# Signal-amplifying Biohybrid Material Circuits for CRISPR/Cas-based single-stranded RNA Detection

**DOI:** 10.1101/2024.06.12.24308852

**Authors:** Hasti Mohsenin, Rosanne Schmachtenberg, Svenja Kemmer, Hanna J. Wagner, Midori Johnston, Sibylle Madlener, Can Dincer, Jens Timmer, Wilfried Weber

**Author notes:** These authors contributed equally (Hasti Mohsenin, Rosanne Schmachtenberg).

## Abstract

The functional integration of biological switches with synthetic building blocks enables the design of modular, stimulus-responsive biohybrid materials. By connecting the individual modules via diffusible signals, information-processing circuits can be designed. Such systems are, however, mostly limited to respond to either small molecules, proteins, or optical input thus limiting the sensing and application scope of the material circuits. Here, we design a highly modular biohybrid material based on CRISPR-Cas13a to translate arbitrary single-stranded RNAs into a biomolecular material response. We exemplify this system by the development of a cascade of communicating materials that can detect the tumor biomarker microRNA miR19b in patient samples or sequences specific for COVID-19. Specificity of the system is further demonstrated by discriminating between input miRNA sequences with single-nucleotide differences. To quantitatively understand information processing in the materials cascade, we developed a mathematical model. The model was used to guide systems design for enhancing signal amplification functionality of the overall materials system. The newly designed modular materials can be used to interface desired RNA input with stimulus-responsive and information-processing materials for building point-of-care suitable sensors as well as multi-input diagnostic systems with integrated data processing and interpretation.

## 1. Introduction

The integration of synthetic biology with materials sciences bears a huge potential for the development of innovative materials systems capable of sensing and responding to different external triggers that find their applications in diagnostics,[1] drug delivery,[2] tissue engineering[3] and personalized medicine[4]. Research at the interface of these two fields started in the early 2000s[5] with the first DNA origamis,[6] a self-ordering system based on engineered bacteriophages,[7] and biomaterials patterned through a genetically manipulated bacterial co-culture.[8] Within the following 20 years, the field of smart biohybrid materials appeared bringing along materials with stimuli-responsive properties, either directly through synthetic biological building blocks,[1, 4, 9] or by integration of engineered living organisms.[10]

The responsiveness to stimuli makes such intelligent materials ideally suited for detection purposes. Materials designed for analytical applications can, for instance, be employed in the detection of environmental pollutants or clinical biomarkers. The most studied types of analytes for synthetic biology-based materials are proteins,[11] nucleic acids,[12] and various small molecular compounds.[9, 13] However, few materials are dedicated to RNA detection, possibly due to the comparably low stability of many RNA targets. In addition to that, the typically low abundance often requires a preamplification and RNA isolation step which increases the intricacy of the system. Examples of RNA sensing materials include various nanomaterials, mainly based on gold or silver nanoparticles, magnetic nanoparticles, quantum dots, or carbon nanomaterials (e.g. graphene oxide). Here, the materials serve not only as support for nucleic acid probes but also function as signal enhancer.[13i, 14] High specificities and sensitivities with detection limits in the femtomolar to attomolar range have been achieved with those detection materials, highlighting their importance in detecting RNA biomarkers, particularly at extremely low concentrations. Another important group of RNA sensors are paper-based systems. They utilize for instance toehold switches or CRISPR/Cas for the detection of mRNAs and viral genomes.[12c, 12d, 16] These systems offer great accessibility for point-of-care use due to their simple handling and low costs. Despite their exceptional performance, most of the nanomaterials and paper-based sensors for RNA detection employ a prior nucleic acid amplification step which increases the system’s complexity and might compomise selectivity.

Most developed sensing materials respond to the presence of a trigger molecule with a change in the material stiffness or the release of a material-bound compound. For instance, there are functionalized acrylamide hydrogels that swell upon the addition of free antigen,[11a, 13c] or release entrapped pharmaceuticals in response to the presence of an antibiotic.[9] In another system, a gel forms through hybridization of an aptamer and DNA-polyacrylamide disintegrates in the presence of cocaine.[13e]

In contrast to those straightforward sensing materials that translate an input signal to a certain output, information processing materials treat input signals in a more sophisticated way. The latter can, for instance, use logic gates or combinations thereof to program a material’s response to a trigger. The key enabling technologies for these materials are synthetic biology and the integration of biological building blocks into materials, as they offer a great range of possibilities for information manipulation such as integration,[17] filtering,[18] memory,[19] and signal modulation.[20] There are several such synthetic biology-based materials that act as simple logic gates such as AND, OR, YES and NOT gates.[13i, 21] To expand the range of possible behaviours, such gates were combined, resulting in for example NOR, INHIBIT, IF-THEN gates or a collection of hierarchical systems responding differently to each combination of three different inputs.[21c] By combining two OR gates, a materials system based on functionalized crosslinked agarose was created that acts as 4 input 2 output binary encoder by processing protease signals.[22] In another work, a binary decoder for the detection of 2 inputs was generated to provide 4 distinct outputs.[13h] Moreover, other motifs are employed in responsive materials. There are, for example, sensing materials incorporating feedforward and feedback loops for signal amplification,[23] and a counting material that generates different outputs as a function of the number of detected light pulses.[24]

Sensing materials can also be created using components of a CRISPR/Cas system for the detection of nucleic acids. For DNAs, the Cas12a effector, an activtable nuclease, is especially well suited for this type of application since it is characterized by flexible programmability, sequence-specificity, and intrinsic signal amplification.[12b, 25] An interesting example for a DNA detection material is shown by combining hydrogels with a CRISPR/Cas system.[12b] In these hydrogels, the Cas12a effector releases immobilized molecules or entrapped cargo once activated by a specific double-stranded DNA (dsDNA).[12b] Another relevant member of the CRISPR/Cas effector class 2 is Cas13a. It forms a complex with a crRNA (CRISPR RNA), a single-stranded RNA (ssRNA) that mediates sequence-specific hybridization with a complementary target ssRNA. Upon target recognition, the complex transitions to a catalytically active state, in which Cas13a degrades ssRNA in a non-specific manner. This collateral cleavage activity results in the cleavage of numerous RNA molecules per activated Cas13a complex.[16b, 26] This property was leveraged in nucleic acid detection systems like SHERLOCK where isothermal amplification and reverse transcription is combined with Cas13a-mediated reporter cleavage.[16b]

In this work, we introduce, the all-in-one concept of a highly modular material circuit, based on magnetic polymers and synthetic biology building blocks, for specific target-amplification free CRISPR-powered RNA sensing. The synthetic network used in combination with Cas13a allows highly specific conversion and amplification of ssRNA signals without the need for prior reverse transcription or isothermal amplification. Additionally, a protease-based material module was also incorporated to further enhance the CRISPR/Cas13a-based signals. Simple fluorescence measurements could finally provide insight into the presence or absence of a certain ssRNA. High modularity is given at several levels: (i) Target specificity is customizable through adapting the crRNA, (ii) the subsequent signal amplification is tunable and (iii) the output molecule is interchangeable with another fluorescent protein, dye or reporter enzyme. This newly designed sensing system highlights the potential of combining Cas13a with material-bound modules, and its application for clinical use.

To develop our RNA sensing, signal amplifying materials circuit, we followed a stepwise approach. After designing a cascade circuit, the three modules of the network were built and characterized. Then, for the proof-of-concept, the entire system was assembled with the biomarker miR19b as target, a microRNA (miRNA) which is dysregulated in children with medulloblastoma. To evaluate its specificity and modularity, and to demonstrate its diagnostic functionality, the assay was tested with other miRNAs and SARS-CoV gene sequences. Finally, our biohybrid diagnostic material was developed further to increase its signal amplification property and with the help of mathematical modelling its behavior could be described and predicted.

## 2. Results and Discussion

### 2.1. Design of ssRNA Detecting Synthetic Biological Material Modules

Here, we propose a design concept for the detection of ssRNAs via CRISPR/Cas13a technology combined with signal amplification based on inter-material communication. For this aim, we assembled biological molecules on material frameworks in a cascade design, in which an input (IN) is detected by a first module (A), then transmitted by a second module (B) to a third module (C) creating an output signal (OUT) (**Figure 1A**).

**Figure 1.**
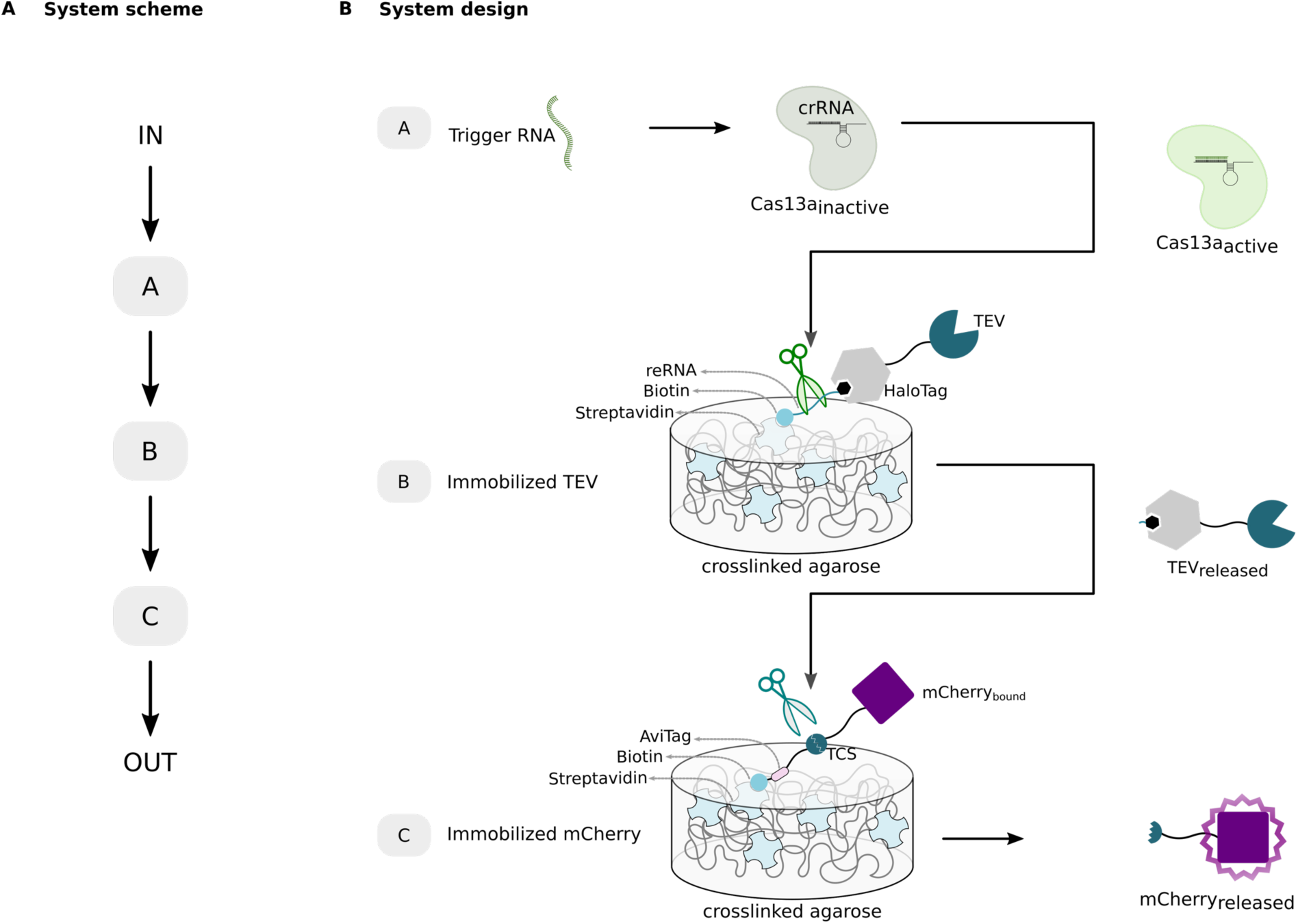
Design of a ssRNA detecting materials system via CRISPR/Cas technology. **A**) System scheme describing the cascade design of our approach built from three consecutive modules allowing signal transmission and the conversion of an input signal (IN) to an output (OUT). **B)** System design. The Cas module is a complex of crRNA and Cas13a that is activated upon binding of the trigger RNA to the crRNA. In the TEV module, reporter RNA (reRNA) is bound via its biotinylated 3’ end to streptavidin on the surface of magnetic crosslinked agarose. On its 5’ end it is covalently bound to HaloTag-TEV via a HaloTag-ligand. For the mCherry module, mCherry is genetically fused to an AviTag via a linker with TEV cleavage site (TCS). The AviTag is biotinylated and thus, immobilizes the protein on streptavidin-coated magnetic crosslinked agarose. Presence of the trigger RNA activates Cas13a, that collaterally cleaves the reRNA, resulting in the release of TEV. TEV subsequently cleaves at the TCS in the linker and releases mCherry from its solid support to enable the readout of output fluorescence signal.

To realize this, a set of biological building blocks was designed and developed, and subsequently compiled together in series to measure a defined ssRNA as the input, transmit the signal between different material modules, and generate a conveniently measurable output signal. As depicted in **Figure 1B**, our system works as follows: Cas13a/crRNA complex detects its trigger ssRNA and gets activated upon binding. When activated, Cas13a shows a collateral RNA cleavage activity and is able to cleave non-specifically any ssRNA in its surrounding. To propagate the information to another module, the second module (TEV module) was designed with an ssRNA as cleavable linker for an immobilized functional protein. This cleavable ssRNA linker serves as reporter RNA (reRNA) and is 5’– and 3’-terminally functionalized with a HaloTag-ligand and a biotin, respectively. The HaloTag-ligand permits its covalent coupling to a HaloTag, which is genetically fused to the tobacco etch virus protease (TEV). The biotinylated end of the reRNA binds with high affinity (K_d_ ∼ 10^-15^ M)[27] to streptavidin on magnetic crosslinked agarose. Thus, the reRNA molecule permits immobilization of TEV on its streptavidin-functionalized support, as well as the release of TEV to the supernatant in the presence of activated Cas13a. The third module (mCherry module) of the system was designed to connect the TEV cleavage activity to the generation of a fluorescence output signal. It contains a linker with a peptide sequence that is specifically cleaved by TEV, the so-called TEV cleavage site (TCS). The mCherry fusion protein consists of an mCherry fluorescent protein genetically fused to a biotinylated AviTag via a peptide linker with TCS. The biotinylation of this protein allows its embedding on streptavidin-functionalized magnetic crosslinked agarose via the biotin-streptavidin interaction. The released TEV would set the mCherry protein free and the fluorescence intensity of the supernatant can be measured as the output. In total, if the trigger RNA is present, activated Cas13a cleaves the reRNA, TEV is released from its material support and would cleave its peptide substrate to free mCherry. Thus, the modules communicate with each other through controlled release of material-bound proteins by cleavage of their respective linkers.

### 2.2. Generation and Characterization of the Material Subsystems

Prior to the assembly of the combined materials system for the detection of ssRNA, two material subsystems were developed and characterized separately (**Figure 2**). For this, the combination of the Cas13a and TEV module was characterized as shown in **Figure 2A**. First, for the Cas module, the Cas13a construct was produced and purified (**Figure S1A**) and cleavage activity was tested (**Figure S1B**). For the TEV module, the HaloTag-fused TEV protease was produced and purified via immobilized metal affinity chromatography (IMAC) and the specific activity determined (**Figure S2**). Subsequently, this protein was immobilized on streptavidin-functionalized magnetic agarose yielding the TEV material module. For characterizing the combination of these two modules, both Cas13a and TEV modules were prepared separately and then combined by transferring a solution containing the Cas13a complex in the well of a microtiter plate with the TEV material module. Our system was established with the miRNA miR19b as target, an important biomarker for medulloblastoma diagnosis. We monitored the activity of the TEV protease in the supernatant at different time points at different miR19b concentrations and deduced the respective TEV concentrations. The concentrations of the released TEV protease increase in a time– and miR19b dose-dependent manner thereby confirming the functionality of the controlled TEV release by this subsystem.

**Figure 2.**
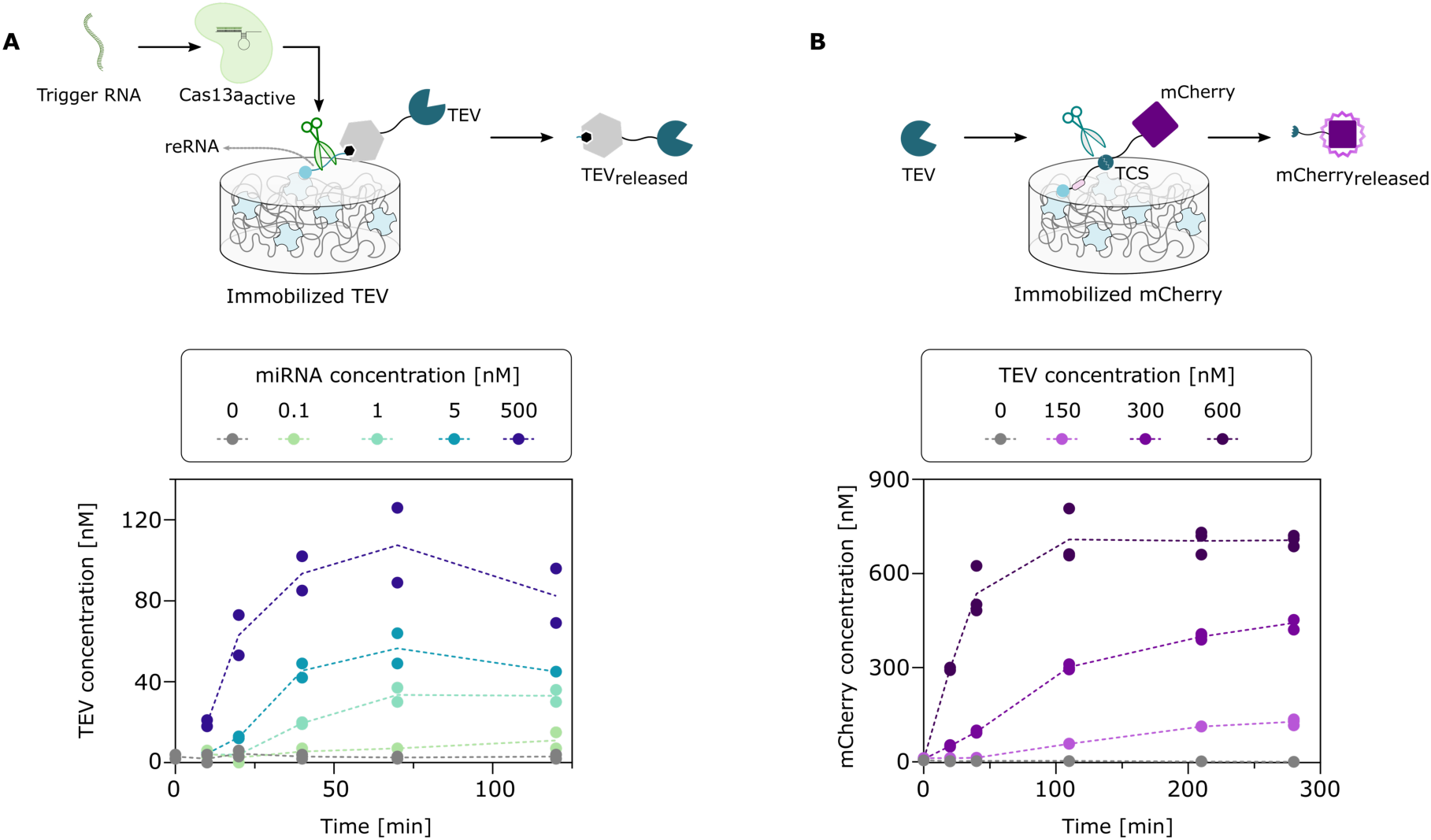
Characterization of Material Subsystems. **A**) The effect of the Cas13a activation on the TEV module. Cas13a is activated by the trigger RNA and cleaves reRNA to release TEV to the supernatant. The release of TEV in the presence of different trigger RNA concentrations is monitored via activity measurements. Experimentally, the TEV module was prepared, the Cas module with crRNA complementary to miR19b was then added as well as miR19b (concentration range: 0 – 500 nM). After different incubation times (0 – 120 min) at 37 °C with gentle mixing, samples of the supernatant were collected and used for TEV activity measurements. For each condition, the activity from a single sample was determined in duplicates. Dotted lines connect the mean values of the replicates. **B)** Characterization of the effect of TEV on the mCherry module. TEV cleaves the TCS of immobilized mCherry and thus releases it to the supernatant. Mobile mCherry is measured via fluorescence measurements. The kinetics of mCherry released with different TEV concentrations were studied. To the freshly prepared mCherry module, free TEV (concentration range: 0 – 600 nM) was added and incubated at 37 °C with gentle mixing. Samples of the supernatant were taken after different incubation times (0 – 280 min) to determine the released mCherry based on fluorescence measurements and a calibration curve (Figure S3B). For each sample, the fluorescence measurement was performed in triplicates. Dotted lines connect the mean values of the replicates.

We then generated the mCherry module, as depicted in **Figure 2B**. mCherry-AviTag was co-expressed with the biotin ligase BirA for *in-vivo* biotinylation and purified by IMAC (**Figure S3**). The biotinylation of the protein construct was validated by a binding test to streptavidin-functionalized crosslinked agarose (**Figure S3C**). Once it was shown to be functional, the protein was immobilized on its material support, and its response to increasing amounts of free TEV construct was monitored over time by profiling mCherry fluorescence in the supernatant (**Figure 2B**).

### 2.3. System Assembly and Modularity Test

Once all the subsystems were shown to be functional, the different modules were assembled together to test the functionality of the cascade. As the presence of all modules in direct physical contact with each other would lead to non-desired interactions and thus, the generation of an output even in the absence of a trigger RNA, we used a custom-made plate stand with embedded bar magnets that is suitable for positioning magnets below each well of a 24-well plate. This installation allows to place each material-bound sub-module above a separate bar magnet (**Figure 3A**). This setup enables communication of the subsystems by the released biomolecules, while preventing direct contact of the material-bound modules.

**Figure 3.**
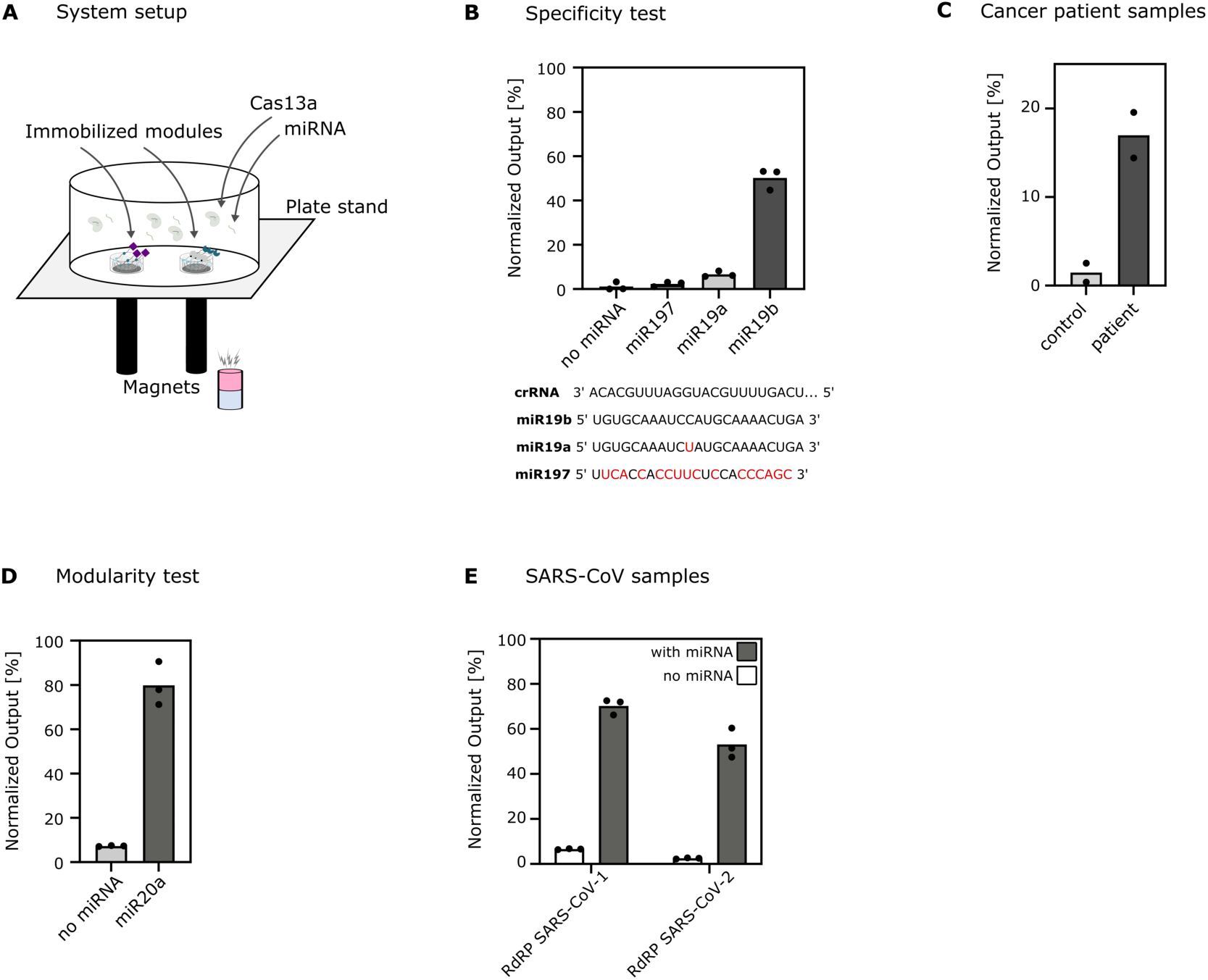
System characterization and proof-of-concept demonstration for RNA diagnostics of different diseases. **A**) System setup. A well of a microtiter plate is placed on top of a plate stand with integrated bar magnets. The modules containing magnetic material (TEV module and mCherry module) are immobilized onto the surface with the help of magnets, whereas the Cas13/crRNA complex and target RNAs are mobile. **B) Specificity test.** The assay was performed with crRNA complementary to the target biomarker miR19b and with reRNA2 sequence (Table 1) as reporter RNA. Comparison of the output signal upon addition of different miRNAs, with full complementarity (miR19b), one mismatch (miR19a), or almost non-complementarity (miR197) with the crRNA used, and in the absence of target miRNA. The raw data was normalized to the fluorescence measurements of the positive and negative controls: supernatant from a well with mCherry module and TEV (1.74 µM) for full release and supernatant from a well with mCherry module, respectively (no release). **C) Cancer diagnostics on medulloblastoma patient sample.** Detection of miR19b in serum sample of a patient suffering from medulloblastoma compared to a control patient serum sample (total RNA concentration of each serum sample: 6 ng µL^-1^; miR19b levels determined by qPCR were 63-fold increased in cancer patient compared to healthy subject). The assay was performed with crRNA complementary to the tumor biomarker miR19b. Data reported as mean, showing duplicate measurements. Values are normalized to the negative (without miR19b) and positive control (500 nM miR19b) measurements. **D) Modularity test.** The assay was performed with crRNA complementary to another target biomarker for medulloblastoma, miR20a, in the presence (10 nM) and in the absence of miR20a with 2-hour incubation. The raw data was normalized to the fluorescence measurements of the positive and negative controls, with (1.56 µM) and without mCherry in assay buffer, respectively. **E) COVID-19 diagnostics.** The Cas13a was equipped with a crRNA either complementary to the RdRP gene of SARS-CoV-1 or the RdRP gene of SARS-CoV-2. The assay was performed with one of those complexes in absence and presence (5 nM) of the corresponding ssRNA RdRP sequence. mCherry fluorescence was measured in the supernatant. For each condition, the fluorescence of the sample was determined in triplicates after a 2-hour incubation. The raw data was normalized to the fluorescence measurements of the positive and negative controls, with (1.56 µM) and without mCherry in assay buffer, respectively. All normalizations were performed using the formula (S-N)/(P-N) with S, P, N as the measurements from the sample, the positive control and the negative control, respectively.

**Table 1.**
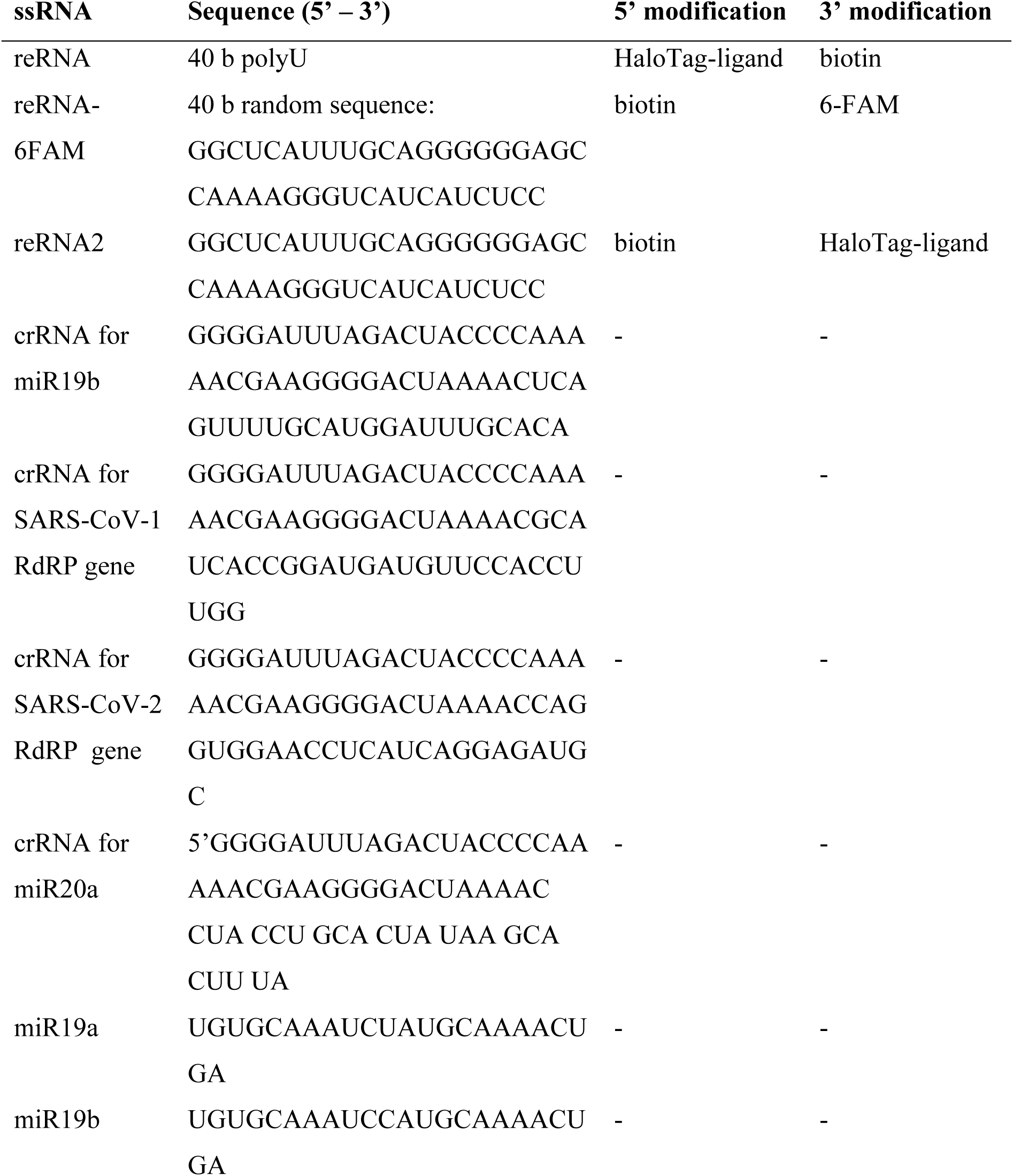
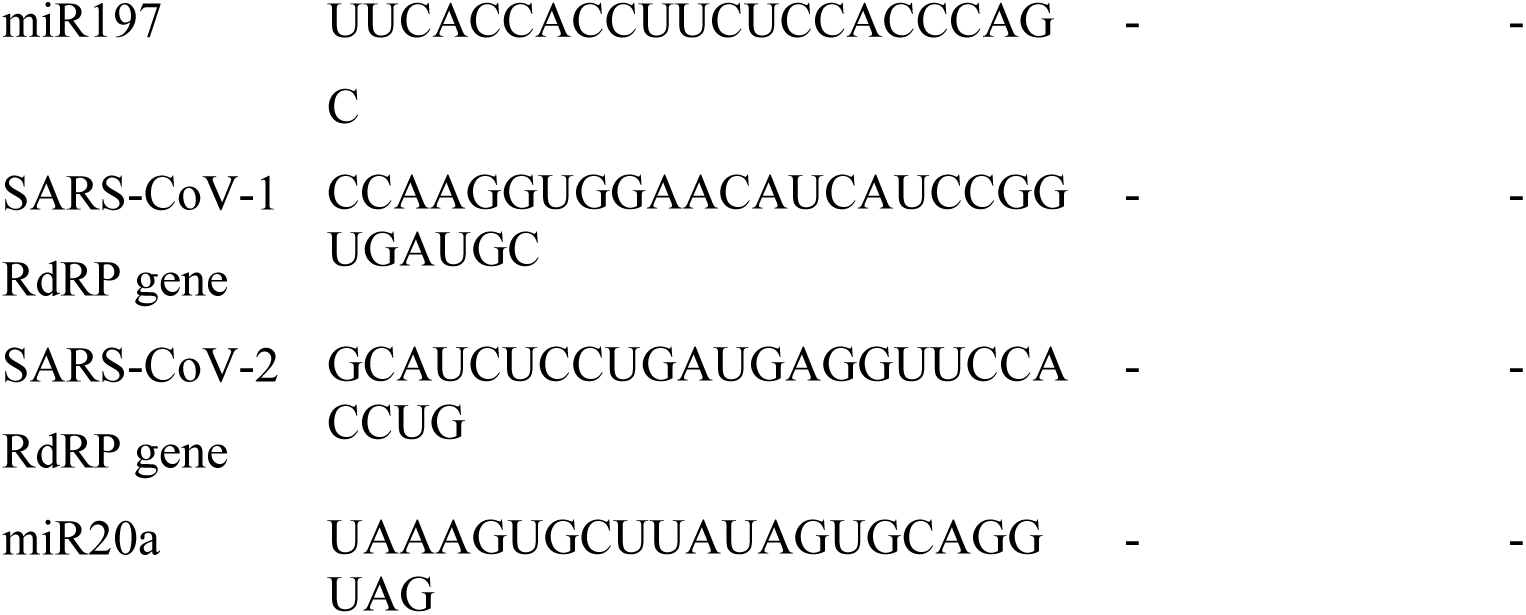
Details of the RNA molecules used in this work.

To test the specificity of the system, we tested the response of the system with three different miRNA sequences using the crRNA targeting miR19b (**Figure 3B**). We tested miR19b which has complete complementarity to the 3’region of the crRNA in comparison to miR19a which has a single mismatch and miR197 which has several mismatches with the crRNA. At 100 nM miRNA input, our system could report a clear output by distinguishing the miRNA of interest from the other miRNAs. The miRNA with only one mismatch (miR19a) showed slightly higher signals compared to no miRNA and miR197 but still marked an obvious signal reduction compared to miR19b. This indicates that the Cas13a complex is activated by molecules with complete complementarity to the 3’region of its crRNA, but not by similar miRNAs. Furthermore, we tested if the system could detect the biomarker miR19b in serum samples of patients suffering from medulloblastoma (**Figure 3C**). For this, we assembled the system as decribed above and added the serum samples with a total RNA concentration of 6 ng µL^-1^ (serum control pool, and medulloblastoma patient sample). After a 4-hour incubation, we measured the fluorescence intensity of duplicates and could observe a difference between the control and the patient sample. This indicates that the system exhibits a sensitivity in a disease-relevant range. To evaluate the modularity of the system, we first assembled all three modules as before, with the modification that the Cas13a/crRNA complex was programmed to detect another RNA target by exchanging the sequence of its crRNA. It was tested if the system could detect the tumour specific miRNA miR20a by performing the detection assay with miR20a-targeting crRNA in the absence and presence (10 nM) of miR20a and monitored the mCherry fluorescence intensity after 2 h (**Figure 3D**). Moreover, it was further explored if the detection of characteristic viral RNA sequences could be detected. For this, the genes for RNA-dependent RNA polymerases (RdRP) from the two viruses, severe acute respiratory syndrome coronavirus (SARS-CoV)-1 and SARS-CoV-2, were chosen (**Figure 3E**). For this purpose, the 3’ region of the crRNA was changed to the reverse complement of each RdRP gene sequence, and the response of the system was measured after 2 h in the presence of the respective ssRNA RdRP gene (5 nM), or in their absence. The output is reported in % as the fluorescence intensity was normalized to the theoretical maximum release (1.56 µM). The system was able to successfully detect the specific SARS-CoV RNA sequences.

With these tests, we showed that by combining the individual modules to a simple network, we can achieve decent analytics. The system is highly specific to the target sequence and able to distinguish between tumor and control patient samples without any target pre-amplification. Furthermore, the system was shown to be highly modular, meaning that by adapting only one component of the system, the crRNA, we can adjust the system to detect a specific clinically relevant miRNA of interest as well as viral ssRNA targets.

### 2.4. System Characterization by a Mathematical Model Towards Improved Signal Amplification

In the setup described above, the TEV module shows a slight signal amplification towards the mCherry module. The intrinsic amplification functionality of the TEV protease, as building block of the TEV module formed the perfect basis for such a signal amplification. However, in the original system design, mCherry with TCS, the substrate of TEV, was not available in excess, thereby limiting the potential of amplification. In order to overcome this issue, we switched the crosslinked material used for mCherry module from magnetic agarose to a cellulose-based magnetic material that has more convenient capturing and release properties for the mCherry protein used (**Figure 4A, Figure S3C**). This material consists of larger particles (30 – 50 µm *versus* 0.6 µm), which may facilitate diffusion of the proteins between the particles and immobilization of the magnetic particles by the magnets.

**Figure 4.**
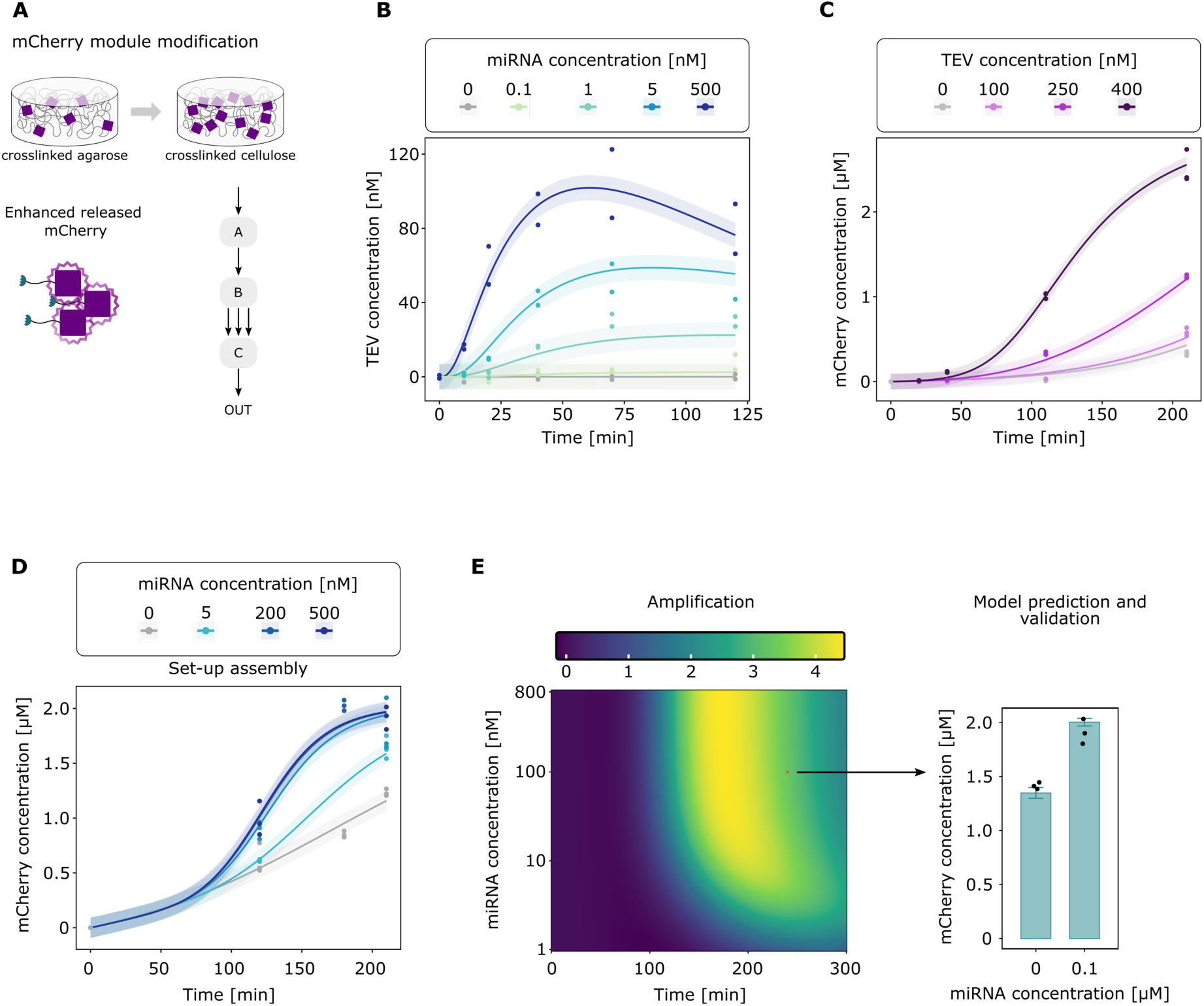
Mathematical model-based optimization of the developed signal-amplifying biohybrid material circuit. **A**) mCherry module modification. mCherry was immobilized on another magnetic material with higher binding capacity (macroporous cellulose instead of crosslinked agarose). Higher amounts can thus be introduced in the system allowing a stronger signal amplification. **B)** Modelling the interaction of Cas13/crRNA complex and TEV module. Fit of the mathematical model and experimental data for the time-resolved release of TEV protease induced by different concentrations of trigger RNA (same data as shown in Figure 2A). Here, the Cas13a complex was equipped with a crRNA complementary to the miR19b sequence and was incubated together with the TEV module and miR19b (0 – 500 nM). **C)** Modelling TEV-mediated release of mCherry from the mCherry module. The modified mCherry module described in A) was incubated in the presence of mobile TEV protease (0 – 400 nM). Samples from the supernatant were taken after different times to measure the fluorescence intensity from released mCherry in triplicates. **D)** Modelling the whole system including all three modules. The Cas13, TEV and mCherry modules were incubated together with different concentrations of the trigger miR19b. At different time points, samples were taken for fluorescence measurements. **E)** Prediction and validation of the mathematical model. (left) Heatmap showing the prediced amplification behavior of the system as a function of miRNA concentration and time. The amplification is the factor by which the amount of released mCherry is higher than the amount of TEV. (right) Validation of model predictions by measurement for two different conditions after 4-hour incubation. The model predictions are shown in the bar chart with uncertainties whereas the experimental data is represented as black dots in the same plot**. B) –D)** Fits of the mathematical model are indicated as solid lines along with the shaded error bands corresponding to one standard deviation, whereas dots correspond to experimental data.

To disentangle the interaction between the individual modules, characterize the combined system, predict the system’s output to different RNA concentrations and identify promising working conditions, we established a quantitative mathematical model. This model based on ordinary differential equations (ODEs) was first calibrated on the experimental data monitoring the response of each individual subsystem to their respective inputs. This procedure is explained in detail in the Supporting Information. **Figure 4B** and **4C** show the calibrated model fit along with the experimental data monitoring the time-resolved release of TEV and mCherry in response to the addition of different concentrations of miR19b and TEV, respectively. When comparing the performance of the mCherry release by TEV obtained here (Figure 4C) with the previous version (Figure 2B), indicates that the modification of the mCherry module indeed increased signal amplification. Whereas, before there was no or only slight amplification by TEV (approximately 700 nM and 300 nM mCherry released by 600 nM and 300 nM TEV after 2 h), the improved system the amplification is stronger (> 1 µM mCherry released by 400 nM TEV).

The combination of the two subsystems was tested, as shown in **Figure 4D**, with the curves corresponding to the fits of the mathematical model and the dots corresponding to the experimental data. The condition with no miRNA shows a leakiness which indicates that the sensitivity of the system decreases for too long incubation times. However, this condition could be captured by the mathematical model after inclusion of an additional reaction for the leaky release in the combined system. Once calibrated, we used the mathematical model to predict the amplification of the TEV module as function of time and miR19b concentration (**Figure 4E**), where the amplification was defined as ratio of the concentration of free mCherry after subtracting the unspecific leaky release from the negative control to the maximum released TEV concentration for the respective condition. To validate the model, we performed validation measurements for a condition with intermediate amplification behavior: 100 nM miR19b at 4-hour incubation, indicated as red cross on the heatmap in Figure 4E. As shown in the bar chart in **Figure 4E**, our predictions aligned very well with the experimental data, thereby validating our mathematical model. This result shows that the model can indeed be utilized as predictive instrument. Furthermore, it may be applied as design tool to help efficiently find optimal assay conditions and predict the system’s performance under conditions not experimentally tested.

## 3. Conclusion

In this study, we applied engineering concepts to synthetic biology-driven material modules to design and develop a signal-amplifying biohybrid material circuit for RNA diagnostics. Our approach is highly modular, where we can adapt only one component of the system, namely the crRNA, and customize our platform to detect an RNA of interest, for example clinically relevant miRNAs or viral RNAs. Such ssRNAs are indicators for many diseases and have great potential as biomarkers for medical diagnostics. The system was characterized and optimized for output performance, assisted by a mathematical model without the need for expensive instruments. We confirmed the introduced concept by performing validation experiments that could reproduce model-based predictions of the signal amplification behavior of the system.

Another special feature of our system is its high degree of modularity, which makes it customizable at several levels: i) The output could be altered by replacing mCherry by another fluorescent protein or dye. Alternatively, the product of an enzymatic reaction that is rendered possible upon protease cleavage could be measured.[28] Thus, the evaluation of a test result would be possible not only through fluorescence, but also via absorbance, luminescence or even electrochemical [29] signal readouts. ii) Our biohybrid sensing circuit can be easily programmed to other targets, all kinds of ssRNAs. Besides simply exchanging the crRNA in the Cas13a complex to switch to another ssRNA target as shown in this study, the spectrum of potential targets could be further extended. For example, Cas12a in combination with a ssDNA linker would create a dsDNA sensing material. In addition to that, leveraging new CRISPR/Cas-based strategies, for instance, in combination with aptamers and antibodies, the detection of various non-nucleic acid targets might also be possible (such as metal ions, antibiotics, proteins).[30] iii) By tuning the intermediate step of the system’s cascade, the kinetics and speed of the signal transmission to the output module can be adjusted. This could be done by choosing from a large pool of sequence-specific proteases.[31] The maximum amplification factor could also be altered by changing the ratio of total protease amount to output molecules in the system.

The concept of CRISPR-powered biohybrid materials systems could be integrated with other technologies, including lateral flow assays or electrochemical sensing devices, and extended to a range of other sensing applications in diagnostics by detecting disease-related biomolecules, or in pharmacokinetics to monitor and optimize drug therapies. Moreover, it might be used in agriculture to assess soil quality and plant health, or to monitor food quality and safety. Biotechnological industry could benefit from it for the surveillance of bioprocesses. Finally, it highlights the potential for developing other sensing material networks for diagnostic applications and beyond.

The advantage of the detection biohybrid materials circuits developed here is that one single type of measurement and assay setup could be used for various different analytes. The underlying principle introduced in this study showed the cruciality of combining several areas of science, from molecular biology and protein engineering to mathematical modelling, materials sciences and engineering. And it highlights its potential for developing sensing materials networks for applications in the field of molecular detection systems.

## Experimental Section/Methods

### Nucleic acids

The expression vectors for TEV and mCherry constructs (Table S2) were generated via PCR and Gibson Assembly. For Cas13a, the expression vector p2CT-His-MBP-Lbu_C2c2_WT was purchased from Addgene (Plasmid #83482). All ssRNAs were purchased from Biomers GmbH (**Table 1**).

### Protein production and analysis

BL21(DE3) pLysS *E. coli* (Invitrogen) were transformed with the respective expression plasmid and grown in LB medium supplemented with ampicillin (100 µg mL^-1^) and chloramphenicol (36 µg mL^-1^) at 150 rpm and 37 °C until the OD_600nm_ reached 0.6. After induction of expression by the addition of isopropyl-β-D-1-thiogalactopyranoside (IPTG, 1 mM), the cells were further incubated at 150 rpm. For Cas13a (Lbu Cas13a of *Leptotrichia buccalis*), the production was performed at 16 °C overnight, for TEV at 30 °C for 3 h, and for mCherry at 37 °C for 2 h. For the production of biotinylated proteins, biotin (50 µM) was added at the induction point for mCherry cultures. Cells were harvested by centrifugation at 6,000 g for 10 min at 4 °C and subsequently resuspended in Ni-Lysis buffer (50 mM NaH_2_PO_4_, 300 mM NaCl, 10 mM imidazole, pH 8.0), flash frozen in liquid nitrogen and stored at – 80 °C.

The proteins were purified via IMAC performed according to Wagner et al. 2019[23b] except for the lysis of cells expressing Cas13a which was achieved through liquid homogenization with a laboratory homogenizer APV-2000 (SPXFLOW) instead of sonication. Each protein was stored in elution buffer (50 mM NaH_2_PO_4_, 300 mM NaCl, 250 mM imidazole, pH 8.0). TEV eluate was supplemented with β-mercaptoethanol (10 mM) and Cas13a eluate with tris(2– carboxyethyl)phosphine (TCEP, 1 mM). Purified Cas13a and TEV were stored at – 80 °C with glycerol (15 % v/v) whereas mCherry was stored at 4 °C.

The concentrations of the individual proteins were determined by analyzing SDS gels with ImageJ. In brief, a dilution series of bovine serum albumin (BSA) as standard (7.8 – 500 µg mL^-1^) and the samples were loaded on a gel. From the pixel intensities in the concerned region of the respective lane, the concentrations were deduced based on the calibration curve from the BSA dilutions.

The proteolytic activity of TEV was determined with the SensoLyte^®^ 520 TEV Protease Assay Kit (AnaSpec) according to the manufacturer’s instructions with a total reaction volume of 16 µL in 384-well plates. For data analysis, the slopes of the fluorescence increase due to quencher release were determined, then the value of the minimum slope determined in the whole assay was subtracted from each slope. Based on those corrected slope values, the concentration was determined based on the calibration curve (see Figure S2B). For the determination of mCherry concentrations, the fluorescence was measured (excitation: 575 nm, emission: 620 nm).

### Functionalization of materials

HaloTag-TEV and reRNA were coupled to each other for ≥ 5 h with gentle mixing at 4 °C in assay buffer (AB) (40 mM Tris-HCl, 60 mM NaCl, 6 mM MgCl_2_, pH 7.3) with murine RNase inhibitor (NEB, 1 U µL^-1^) and TCEP (1 mM) with a 5-fold excess of TEV in order to saturate the binding sites of reRNA. The TEV-reRNA was then immobilized on pre-washed (3 times with PBS (1 mL) with Tween20 (0.05 % v/v) and magnetic stand) magnetic crosslinked agarose (MagSi-STA 600 BI, MD21001, Steinbrenner Laborsysteme GmbH) by incubation with gentle mixing overnight at 4 °C. The biotinylated mCherry constructs were diluted in AB with TCEP (1 mM) and then coupled to the magnetic agarose following the same procedure as for reRNA-coupled protease. Functionalized crosslinked agarose was washed 5 times with wash buffer (PBS with TCEP (1 mM), 250 µL). For TEV-reRNA, wash buffer was supplemented with RNase inhibitor (1 U µL^-1^). The washed crosslinked agarose was then resuspended in AB with RNase inhibitor (1 U µL^-1^) and/or TCEP (1 mM). For the modified mCherry module, mCherry-AviTag was coupled to magnetic macroporous cellulose (High Capacity Magne^®^ Streptavidin Beads, V7820, Promega) and TBS instead of PBS was used for all washing steps.

### Detection assay

For the assay, Cas/crRNA complex was assembled by incubating Cas13a (1 μM) with crRNA (0.5 μM) at 37 °C for 30 min in AB. The assay was performed in 24-well plates filled with assay mix (pre-assembled Cas13a/crRNA (100 nM/50 nM), RNase inhibitor (1 U µL^-1^), and TCEP (1 mM) in AB) and two separate spots of magnetic crosslinked agarose, one functionalized with TEV-reRNA and another with mCherry in each well. For the amplifying system mCherry functionalized cellulose was separated into two drops.

To ensure that the two types of functionalized material did not mix, the plate was placed onto a custom-made machined support with integrated bar magnets (Ø 3 mm, height 10 mm) (**Figure 3A**). With this support, underneath each well of a 24-well microtiter plate, three magnets are arranged in a concentric formation with uniform spacing (5.5 mm).

Then, a ssRNA dilution of miR19a, miR19b, miR197, miR20a, SARS-CoV-1 RdRP RNA, or SARS-CoV-2 RdRP RNA in AB was added (final concentrations: 0 – 500 nM) and the plate was sealed with adhesive tape and covered with aluminium foil. The assay plate was incubated at 37 °C with orbital shaking at 25 – 50 rpm. After different incubation times, samples were taken for determination of TEV activity and mCherry fluorescence. The samples were stored at 4 °C until the measurements. For the detection assay with clinical samples, 1 µL samples were added per reaction with 200 µL total volume.

### Characterization of the release kinetics of the individual modules

For the characterization of the TEV module, the detection assay was performed as described above but without the mCherry module. As an output, the enzymatic activity of TEV in the supernatant was monitored.

For the characterization of the mCherry module, the detection assay was performed without the TEV module, Cas13a, and any ssRNA. The release of mCherry was determined after different incubation times in the presence of different concentrations of TEV.

## Supporting Information

Supporting Information is available from the Wiley Online Library or from the author.

## Data Availability

All data produced in the present study are available upon reasonable request to the authors

## Acknowledgments

This work was supported by the European Union (ERC, STEADY, 101053857), the German Research Foundation (Deutsche Forschungsgemeinschaft, DFG) under Germany’s Excellence Strategy –CIBSS, EXC-2189, Project ID: 390939984 and under the Excellence Initiative of the German Federal and State Governments – BIOSS, EXC-294, by the German Ministry of Education and Research (BMBF) through LiSyM Cancer (Grant No. 031L0256G) and in part by the Ministry for Science, Research and Arts of the state of Baden-Württemberg. The authors acknowledge support by the state of Baden-Württemberg through bwHPC. C.D. and M.J. gratefully acknowledge the support by the Bundesministerium für Bildung und Forschung (BMBF, Federal Ministry of Education and Research) under grant number 13GW0493 (MERGE).

## Ethics

Collection of plasma samples from patients was approved by the Ethic commission of the Medical university of Vienna. The title of the Ethic vote is “Bestimmung von Biomarkern in Tumorgewebe, Liquor und Blut bei Patienten mit Hirn-und Rückenmarkstumoren” (No. 1244/2016).

Received: ((will be filled in by the editorial staff))

Revised: ((will be filled in by the editorial staff))

Published online: ((will be filled in by the editorial staff)

## Notes

### Competing Interest Statement

The authors have declared no competing interest.

### Funding Statement

European Union (ERC, STEADY, 101053857)
German Research Foundation (DFG) under Germany's Excellence Strategy CIBSS, EXC2189, Project ID: 390939984
and under the Excellence Initiative of the German Federal and State Governments BIOSS, EXC294,
by the German Ministry of Education and Research (BMBF) through LiSyM Cancer (Grant No. 031L0256G) in part by the Ministry for Science, Research and Arts of the state of Baden-Wuerttemberg
by the state of Baden-Wuerttemberg through bwHPC
Bundesministerium fuer Bildung und Forschung (BMBF) under grant number 13GW0493 (MERGE)

## References

[1] H. J. Wagner, H. Mohsenin, W. Weber, Adv Biochem Eng Biot 2021, 178, 197.

[2] J. Y. Li, D. J. Mooney, Nat Rev Mater 2016, 1 (12).

[3] H. J. Wagner, A. Sprenger, B. Rebmann, W. Weber, Adv Drug Deliver Rev 2016, 105, 77.

[4] B. A. Badeau, C. A. DeForest, Annu Rev Biomed Eng 2019, 21, 241.

[5] T. C. Tang, B. L. An, Y. Y. Huang, S. Vasikaran, Y. Y. Wang, X. Y. Jiang, T. K. Lu, C. Zhong, Nat Rev Mater 2021, 6 (4), 332.

[6] P. W. K. Rothemund, Nature 2006, 440 (7082), 297.

[7] S. W. Lee, C. B. Mao, C. E. Flynn, A. M. Belcher, Science 2002, 296 (5569), 892.

[8] S. Basu, Y. Gerchman, C. H. Collins, F. H. Arnold, R. Weiss, Nature 2005, 434 (7037), 1130.

[9] M. Ehrbar, R. Schoenmakers, E. H. Christen, M. Fussenegger, W. Weber, Nat Mater 2008, 7 (10), 800.

[10a] P. K. R. Tay, P. Q. Nguyen, N. S. Joshi, Acs Synth Biol 2017, 6 (10), 1841

[10b] S. Sankaran, S. F. Zhao, C. Muth, J. Paez, A. del Campo, Adv Sci 2018, 5 (8)

[10c] A. Y. Chen, C. Zhong, T. K. Lu, Acs Synth Biol 2015, 4 (1), 8.

[11a] T. Miyata, N. Asami, T. Uragami, Nature 1999, 399 (6738), 766

[11b] R. H. Wang, Y. B. Li, Biosens Bioelectron 2013, 42, 148

[11c] M. F. Maitz, U. Freudenberg, M. V. Tsurkan, M. Fischer, T. Beyrich, C. Werner, Nat Commun 2013, 4

[11d] C. Geraths, L. Eichstädter, R. J. Gübeli, E. H. Christen, C. Friedrich, W. Weber, J Control Release 2013, 165 (1), 38.

[12a] R. V. Gayet, H. de Puig, M. A. English, L. R. Soenksen, P. Q. Nguyen, A. S. Mao, N. M. Angenent-Mari, J. J. Collins, Nat Protoc 2020, 15 (9), 3030

[12b] M. A. English, L. R. Soenksen, R. V. Gayet, H. de Puig, N. M. Angenent-Mari, A. S. Mao, P. Q. Nguyen, J. J. Collins, Science 2019, 365 (6455), 780

[12c] K. Pardee, A. A. Green, M. K. Takahashi, D. Braff, G. Lambert, J. W. Lee, T. Ferrante, D. Ma, N. Donghia, M. Fan, N. M. Daringer, I. Bosch, D. M. Dudley, D. H. O’Connor, L. Gehrke, J. J. Collins, Cell 2016, 165 (5), 1255

[12d] M. K. Takahashi, X. Tan, A. J. Dy, D. Braff, R. T. Akana, Y. Furuta, N. Donghia, A. Ananthakrishnan, J. J. Collins, Nat Commun 2018, 9.

[13a] E. H. Christen, M. Karlsson, M. M. Kämpf, R. Schoenmakers, R. J. Gübeli, H. M. Wischhusen, C. Friedrich, M. Fussenegger, W. Weber, Adv Funct Mater 2011, 21 (15), 2861

[13b] M. M. Kämpf, E. H. Christen, M. Ehrbar, M. Daoud-El Baba, G. Charpin-El Hamri, M. Fussenegger, W. Weber, Adv Funct Mater 2010, 20 (15), 2534;

[13c] R. J. Gübeli, D. Hövermann, H. Seitz, B. Rebmann, R. G. Schoenmakers, M. Ehrbar, G. Charpin-El Hamri, M. Daoud-El Baba, M. Werner, M. Müller, W. Weber, Adv Funct Mater 2013, 23 (43), 5355

[13d] C. Geraths, M. Daoud-El Baba, G. Charpin-El Hamri, W. Weber, J Control Release 2013, 171 (1), 57

[13e] L. Yan, Z. Zhu, Y. Zou, Y. S. Huang, D. W. Liu, S. S. Jia, D. M. Xu, M. Wu, Y. Zhou, S. Zhou, C. J. Yang, J Am Chem Soc 2013, 135 (10), 3748

[13f] H. H. Yang, H. P. Liu, H. Z. Kang, W. H. Tan, J Am Chem Soc 2008, 130 (20), 6320

[13g] R. D. Liu, Y. S. Huang, Y. L. Ma, S. S. Jia, M. X. Gao, J. X. Li, H. M. Zhang, D. M. Xu, M. Wu, Y. Chen, Z. Zhu, C. Y. Yang, Acs Appl Mater Inter 2015, 7 (12), 6982

[13h] H. Mohsenin, H. J. Wagner, M. Rosenblatt, S. Kemmer, F. Drepper, P. Huesgen, J. Timmer, W. Weber, Adv Mater 2024

[13i] M. P. Nikitin, V. O. Shipunova, S. M. Deyev, P. I. Nikitin, Nat Nanotechnol 2014, 9 (9), 716

[13j] H. C. Ates, H. Mohsenin, C. Wenzel, R. T. Glatz, H. J. Wagner, R. Bruch, N. Hoefflin, S. Spassov, L. Streicher, S. Lozano-Zahonero, B. Flamm, R. Trittler, M. J. Hug, M. Köhn, J. Schmidt, S. Schumann, G. A. Urban, W. Weber, C. Dincer, Adv Mater 2022, 34 (2).

[14a] Y. W. C. Cao, R. C. Jin, C. A. Mirkin, Science 2002, 297 (5586), 1536

[14b] S. Su, J. W. Fan, B. Xue, L. H. Yuwen, X. F. Liu, D. Pan, C. H. Fan, L. H. Wang, Acs Appl Mater Inter 2014, 6 (2), 1152

[14c] N. Ibrahim, N. D. Jamaluddin, L. L. Tan, N. Y. M. Yusof, Sensors-Basel 2021, 21 (15)

[14d] H. Zhou, J. Liu, J. J. Xu, S. S. Zhang, H. Y. Chen, Adv Clin Chem 2019, 91, 31

[14e] M. Tian, M. Qiao, C. C. Shen, F. L. Meng, L. A. Frank, V. V. Krasitskaya, T. J. Wang, X. M. Zhang, R. H. Song, Y. X. Li, J. J. Liu, S. C. Xu, J. H. Wang, Appl Surf Sci 2020, 527.

[15a] D. Voccia, M. Sosnowska, F. Bettazzi, G. Roscigno, E. Fratini, V. De Franciscis, G. Condorelli, R. Chitta, F. D’Souza, W. Kutner, I. Palchetti, Biosens Bioelectron 2017, 87, 1012

[15b] D. G. He, L. Hai, H. Z. Wang, R. Wu, H. W. Li, Analyst 2018, 143 (4), 813.

[16a] K. Pardee, A. A. Green, T. Ferrante, D. E. Cameron, A. DaleyKeyser, P. Yin, J. J. Collins, Cell 2014, 159 (4), 940

[16b] J. S. Gootenberg, O. O. Abudayyeh, J. W. Lee, P. Essletzbichler, A. J. Dy, J. Joung, V. Verdine, N. Donghia, N. M. Daringer, C. A. Freije, C. Myhrvold, R. P. Bhattacharyya, J. Livny, A. Regev, E. V. Koonin, D. T. Hung, P. C. Sabeti, J. J. Collins, F. Zhang, Science 2017, 356 (6336), 438.

[17] J. A. N. Brophy, C. A. Voigt, Nat Methods 2014, 11 (5), 508.

[18] J. R. Rubens, G. Selvaggio, T. K. Lu, Nat Commun 2016, 7.

[19] P. Siuti, J. Yazbek, T. K. Lu, Nat Biotechnol 2013, 31 (5), 448.

[20] A. Prindle, J. Selimkhanov, H. Li, I. Razinkov, L. S. Tsimring, J. Hasty, Nature 2014, 508 (7496), 387.

[21a] P. M. Gawade, J. A. Shadish, B. A. Badeau, C. A. DeForest, Adv Mater 2019, 31 (33)

[21b] M. Ikeda, T. Tanida, T. Yoshii, K. Kurotani, S. Onogi, K. Urayama, I. Hamachi, Nat Chem 2014, 6 (6), 511

[21c] B. A. Badeau, M. P. Comerford, C. K. Arakawa, J. A. Shadish, C. A. DeForest, Nat Chem 2018, 10 (3), 251

[21d] E. R. Ruskowitz, M. P. Comerford, B. A. Badeau, C. A. DeForest, Biomater Sci-Uk 2019, 7 (2), 542.

[22] H. Mohsenin, J. Pacheco, S. Kemmer, H. J. Wagner, N. Höfflin, T. Bergmann, T. Baumann, C. Jerez-Longres, A. Ripp, N. Jork, H. J. Jessen, M. Fussenegger, M. Köhn, J. Timmer, W. Weber, Adv Funct Mater 2023.

[23a] H. J. Wagner, S. Kemmer, R. Engesser, J. Timmer, W. Weber, Adv Sci 2019, 6 (4)

[23b] H. J. Wagner, R. Engesser, K. Ermes, C. Geraths, J. Timmer, W. Weber, Mater Today 2019, 22, 25.

[24] H. M. Beyer, R. Engesser, M. Hörner, J. Koschmieder, P. Beyer, J. Timmer, M. D. Zurbriggen, W. Weber, Adv Mater 2018, 30 (21).

[25] J. S. Chen, E. B. Ma, L. B. Harrington, M. Da Costa, X. R. Tian, J. M. Palefsky, J. A. Doudna, Science 2018, 360 (6387), 436.

[26a] O. O. Abudayyeh, J. S. Gootenberg, S. Konermann, J. Joung, I. M. Slaymaker, D. B. T. Cox, S. Shmakov, K. S. Makarova, E. Semenova, L. Minakhin, K. Severinov, A. Regev, E. S. Lander, E. V. Koonin, F. Zhang, Science 2016, 353 (6299)

[26b] A. East-Seletsky, M. R. O’Connell, S. C. Knight, D. Burstein, J. H. D. Cate, R. Tjian, J. A. Doudna, Nature 2016, 538 (7624), 270

[26c] L. Liu, X. Y. Li, J. Ma, Z. Q. Li, L. L. You, J. Y. Wang, M. Wang, X. Z. Zhang, Y. L. Wang, Cell 2017, 170 (4), 714.

[27] P. C. Weber, D. H. Ohlendorf, J. J. Wendoloski, F. R. Salemme, Science 1989, 243 (4887), 85.

[28a] H. J. Wagner, W. Weber, Molecules 2019, 24 (10)

[28b] T. Azad, A. Tashakor, S. Hosseinkhani, Anal Bioanal Chem 2014, 406 (23), 5541

[28c] R. M. Eglen, R. Singh, Comb Chem High T Scr 2003, 6 (4), 381.

[29a] Y. Sheng, T. H. Zhang, S. H. Zhang, M. Johnston, X. H. Zheng, Y. Y. Shan, T. Liu, Z. N. Huang, F. Y. Qian, Z. H. Xie, Y. R. Ai, H. K. Zhong, T. R. Kuang, C. Dincer, G. A. Urban, J. M. Hu, Biosens Bioelectron 2021, 178

[29b] M. Johnston, H. C. Ates, R. T. Glatz, H. Mohsenin, R. Schmachtenberg, N. Göppert, D. Huzly, G. A. Urban, W. Weber, C. Dincer, Mater Today 2022, 61, 129

[29c] R. Bruch, J. Baaske, C. Chatelle, M. Meirich, S. Madlener, W. Weber, C. Dincer, G. A. Urban, Adv Mater 2019, 31 (51).

[30a] X. K. Cheng, Y. R. Li, J. Kou, D. Liao, W. L. Zhang, L. J. Yin, S. L. Man, L. Ma, Biosens Bioelectron 2022, 215

[30b] W. R. Su, J. R. Li, C. Ji, C. S. Chen, Y. Z. Wang, H. L. Dai, F. Q. Li, P. F. Liu, Nano Res 2023, 16 (7), 9940.

[31a] J. Tözsér, J. E. Tropea, S. Cherry, P. Bagossi, T. D. Copeland, A. Wlodawer, D. S. Waugh, Febs J 2005, 272 (2), 514

[31b] X. Fan, X. Z. Li, Y. Zhou, M. Mei, P. Liu, J. Zhao, W. F. Peng, Z. B. Jiang, S. H. Yang, B. L. Iverson, G. M. Zhang, L. Yi, Acs Chem Biol 2020, 15 (1), 63

[31c] S. Lien, R. Pastor, D. Sutherlin, H. B. Lowman, Protein J 2004, 23 (6), 413.

