## Supporting Information for "Signal-amplifying Biohybrid Material Circuits for CRISPR/Cas-based single-stranded RNA Detection"

**Table S1.** Amino acid sequence of the constructs used in this work. The protein constructs used in this study with their respective plasmids are shown in **Table S1**. The following color code is used: main protein part, affinity tag, solubility tag, hexahistidine tag, protease cleavage site

| **Construct** | **Sequence** |
|---|---|
| $His_6$-MBP-TCS-Cas13a<br>(p2CT-$His_6$-MBP-<br>Lbu_C2c2_WT) | MKSSHHHHHHGSSMKIEEGKLVIWINGDKGYNGLAEVGKKFEKDTGIKV<br>TVEHPDKLEEKFPQVAATGDGPDIIFWAHDRFGGYAQSGLLAEITPDKAF<br>QDKLYPFTWDAVRYNGKLIAYPIAVEALSLIYNKDLLPNPPKTWEEIPAL<br>DKELKAKGKSALMFNLQEPYFTWPLIAADGGYAFKYENGKYDIKDVGV<br>DNAGAKAGLTFLVDLIKNKHMNADTDYSIAEAAFNKGETAMTINGPWA<br>WSNIDTSKVNYGVTVLPTFKGQPSKPFVGVLSAGINAASPNKELAKEFLE<br>NYLLTDEGLEAVNKDKPLGAVALKSYEEELAKDPRIAATMENAQKGEIM<br>PNIPQMSAFWYAVRTAVINAASGRQTVDEALKDAQTNSSSNNNNNNNNN<br>NNLGIEENLYFQSNAMKVTKVGGISHKKYTSEGRLVKSESEENRTDERLS<br>ALLNMRLDMYIKNPSSTETKENQKRIGKLKKFFSNKMVYLKDNTLSLKN<br>GKKENIDREYSETDILESDVRDKKNFAVLKKIYLNENVNSEELEVFRNDIK<br>KKLNKINSLKYSFEKNKANYQKINENNIEKVEGKSKRNIIYDYYRESAKR<br>DAYVSNVKEAFDKLYKEEDIAKLVLEIENLTKLEKYKIREFYHEIIGRKND<br>KENFAKIIYEEIQNVNNMKELIEKVPDMSELKKSQVFYKYYLDKEELNDK<br>NIKYAFCHFVEIEMSQLLKNYVYKRLSNISNDKIKRIFEYQNLKKLIENKL<br>LNKLDTYVRNCGKYNYYLQDGEIATSDFIARNRQNEAFLRNIIGVSSVAY<br>FSLRNILETENENDITGRMRGKTVKNNKGEEKYVSGEVDKIYNENKKNE<br>VKENLKMFYSYDFNMDNKNEIEDFFANIDEAISSIRHGIVHFNLELEGKDI<br>FAFKNIAPSEISKKMFQNEINEKKLKLKIFRQLNSANVFRYLEKYKILNYL<br>KRTRFEFVNKNIPFVPSFTKLYSRIDDLKNSLGIYWKTPKTNDDNKTKEIID<br>AQIYLLKNIYYGEFLNYFMSNNGNFFEISKEIIELNKNDKRNLKTGFYKLQ<br>KFEDIQEKIPKEYLANIQSLYMINAGNQDEEEKDTYIDFIQKIFLKGFMTYL<br>ANNGRLSLIYIGSDEETNTSLAEKKQEFDKFLKKYEQNNNIKIPYEINEFLR |

| | |
|---|---|
| | EIKLGNILKYTERLNMFYLILKLLNHKELTNLKGSLEKYQSANKEEAFSD<br>QLELINLLNLDNNRVTEDFELEADEIGKFLDFNGNKVKDNKELKKFDTNK<br>IYFDGENIIKHRAFYNIKKYGMLNLLEKIADKAGYKISIEELKKYSNKKNEI<br>EKNHKMQENLHRKYARPRKDEKFTDEDYESYKQAIENIEEYTHLKNKVE<br>FNELNLLQGLLLRILHRLVGYTSIWERDLRFRLKGEFPENQYIEEIFNFENK<br>KNVKYKGGQIVEKYIKFYKELHQNDEVKINKYSSANIKVLKQEKKDLYIR<br>NYIAHFNYIPHAEISLLEVLENLRKLLSYDRKLKNAVMKSVVDILKEYGF<br>VATFKIGADKKIGIQTLESEKIVHLKNLKKKKLMTDRNSEELCKLVKIMFE<br>YKMEEKKSEN |
| HaloTag-TEV-$His_6$ (pHM300) | MEIGTGFPFDPHYVEVLGERMHYVDVGPRDGTPVLFLHGNPTSSYVWRN<br>IIPHVAPTHRCIAPDLIGMGKSDKPDLGYFFDDHVRFMDAFIEALGLEEVV<br>LVIHDWGSALGFHWAKRNPERVKGIAFMEFIRPIPTWDEWPEFARETFQA<br>FRTTDVGRKLIIDQNVFIEGTLPMGVVRPLTEVEMDHYREPFLNPVDREPL<br>WRFPNELPIAGEPANIVALVEEYMDWLHQSPVPKLLFWGTPGVLIPPAEA<br>ARLAKSLPNCKAVDIGPGLNLLQEDNPDLIGSEIARWLSTLEISGHGGGSG<br>GGSDEVDGGGSGGGSGESLFKGPRDYNPISSTICHLTNESDGHTTSLYGIG<br>FGPFIITNKHLFRRNNGTLLVQSLHGVFKVKNTTTLQQHLIDGRDMIIIRM<br>PKDFPPFPQKLKFREPQREERICLVTTNFQTKSMSSMVSDTSCTFPSSDGIF<br>WKHWIQTKDGQCGSPLVSTRDGFIVGIHSASNFTNTNNYFTSVPKNFMEL<br>LTNQEAQQWVSGWRLNADSVLWGGHKVFMVKPEEPFQPVKEATQLMN<br>RRRSGGGSHHHHHH |
| mCherry-TCS-AviTag-$His_6$ (pHM303) | MVSKGEEDNMAIIKEFMRFKVHMEGSVNGHEFEIEGEGEGRPYEGTQTA<br>KLKVTKGGPLPFAWDILSPQFMYGSKAYVKHPADIPDYLKLSFPEGFKWE<br>RVMNFEDGGVVTVTQDSSLQDGEFIYKVKLRGTNFPSDGPVMQKKTMG<br>WEASSERMYPEDGALKGEIKQRLKLKDGGHYDAEVKTTYKAKKPVQLP<br>GAYNVNIKLDITSHNEDYTIVEQYERAEGRHSTGGMDELYKSGGGSGGG<br>GENLYFQSGGGPAGEASSIPNREGKPIPNPLLGLGSTRTGEFSLSTPPTPSTP<br>PTGLNDIFEAQKIEWHEHHHHHH |

**Table S2.** Expression vectors of this work.

| Vector | Description |
|---|---|
| pHM300 | $P_{T7}$ HaloTag-TEV-$His_6$, $P_{\beta\text{-lac}}$ $amp^R$ |
| pHM303 | $P_{T7}$ mCherry-TCS-AviTag-$His_6$, $P_{T7}$ *birA*, $P_{lac}$ *lacI*, $P_{\beta\text{-lac}}$ $amp^R$ |

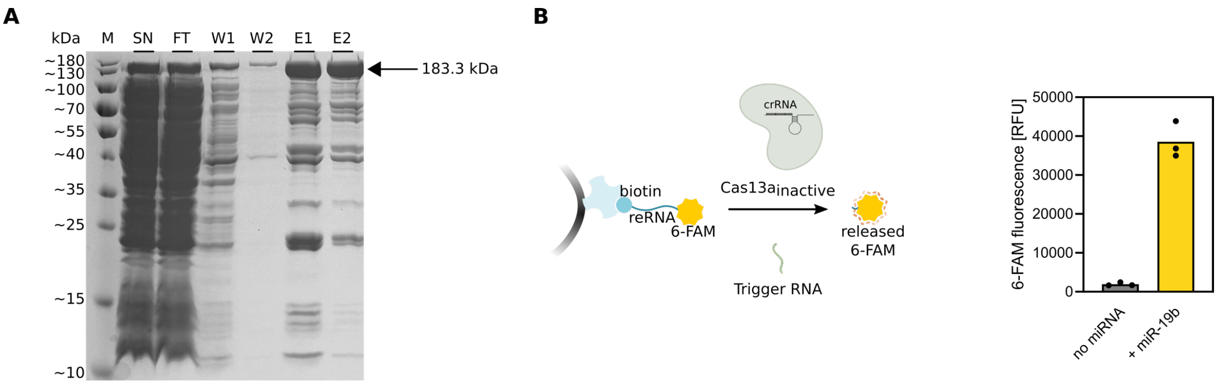


**Figure S1. Characterization of Cas13a protein**. **A)** SDS-PAGE analysis of lbuCas13a (p2CT-His6-MBP-Lbu_C2c2_WT) production in *E. coli* and immobilized metal affinity chromatography (IMAC)on a acrylamide gel (12 % w/v). Expected size of 183.3 kDa. M, marker; SN, supernatant; FT, flow-through; W1, wash fraction 1; W2, wash fraction 2; E1, elution; E2, eluate of the second round of purification. For the second round of purification, cell lysate as well as the flow through from the first round were applied onto the column. The purification was then performed according to the protocol. Finally, both eluates were combined. **B)** Biotinylated RNA functionalized with 6-FAM (reRNA-6FAM) coupled to streptavidin-functionalized magnetic crosslinked agarose incubated with the Cas13a/crRNA complex in the absence and presence (0.75 nM) of miR19b. For this, streptavidin-coated magnetic agarose (MagSi-STA 600, MD16001, Steinbrenner Laborsysteme GmbH) were first washed 5 times with PBS with Tween20 (0.05 % v/v) and then resuspended in wash buffer. ReRNA-6FAM (25 pmol per 100 µg beads) was coupled to the beads in the presence of RNase inhibitor (1 U $\mu L^{-1}$) for 1 h at RT with 600 rpm. For Cas complex formation, Cas13a (1 µM) was incubated with crRNA for miR19b (0.5 µM) for 30 min at 37 °C in AB with RNase inhibitor (1 U $\mu L^{-1}$) and TCEP (1 mM). The beads were incubated for another 2 h at 37 °C and occasional mixing at 15 rpm with pre-assembled Cas13a/crRNA and miR19b (0 or 1 nM). The fluorescence of 6-FAM in the supernatant was measured in triplicates (excitation: 492 nm, emission: 517 nm).

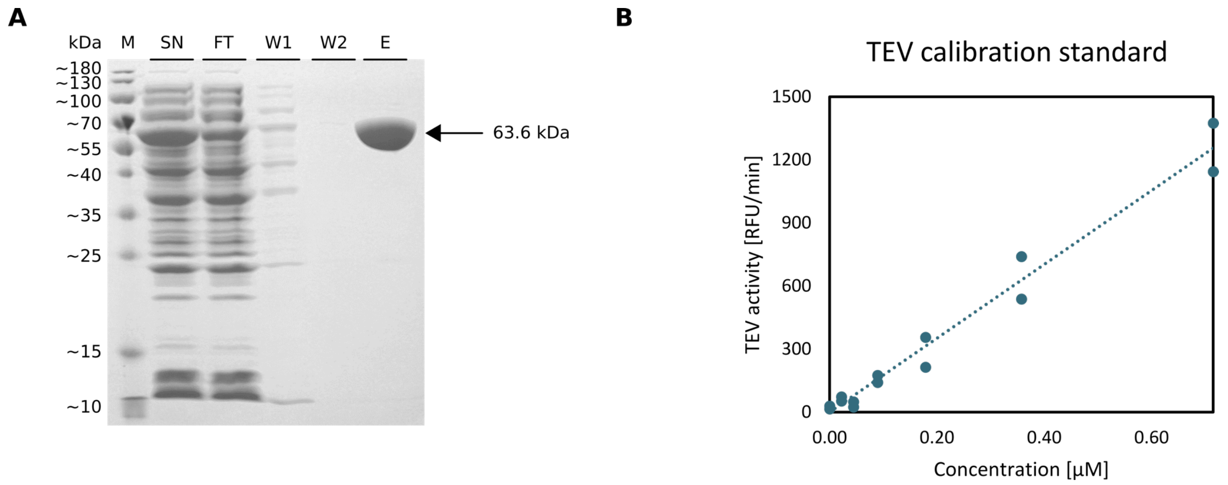


**Figure S2. Characterization of TEV protein. A)** SDS-PAGE analysis of TEV construct (pHM300: HaloTag-TEV-$His_6$) produced in *E. coli* and IMAC on a acrylamide gel (12 % w/v). Expected size of 63.6 kDa. SN, supernatant; FT, flow-through; W1, wash fraction 1; W2, wash fraction 2; E, elution; M, marker. **B)** TEV catalytic activities determined from the slope in the linear phase of the curves obtained upon addition of substrate to TEV protease and fluorescence measurements of 5-FAM (excitation: 490 nm, emission: 520 nm). The specific activity of this enzyme was determined to be 1756.29 U, where we defined the enzyme unit 1 U as the increase in measured fluorescence of 5-FAM in relative fluorescence units per minute and per µM of enzyme in the tested sample.

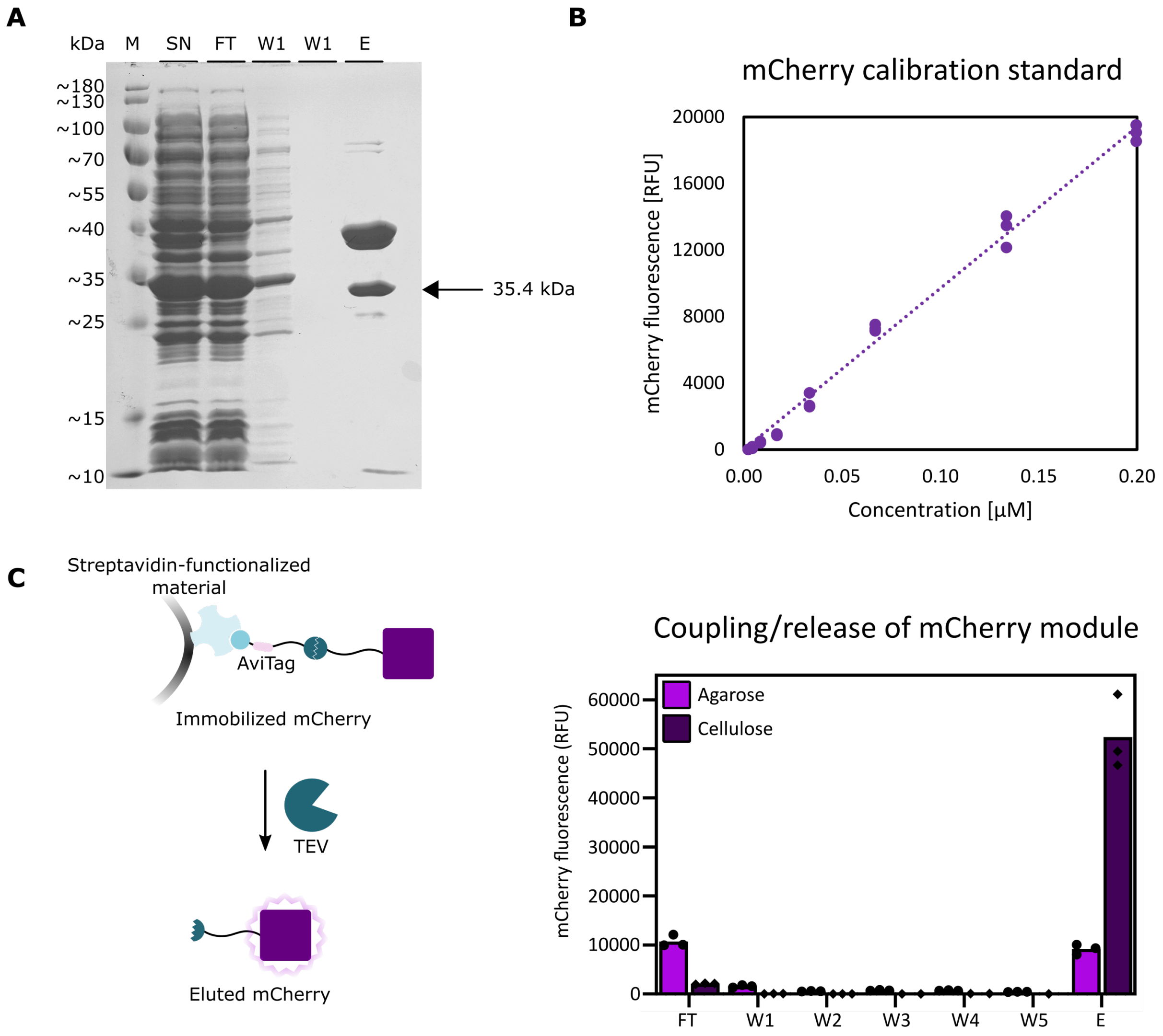


**Figure S3. Characterization of mCherry protein with TEV cleavage site (TCS). A)** SDS-PAGE analysis of mCherry construct (pHM303: mCherry-TCS-AviTag-$His_6$) production in *E. coli* and IMAC on a acrylamide gel (12 % w/v). Expected size of 35.4 kDa. SN, supernatant; FT, flow-through; W1, wash fraction 1; W2, wash fraction 2; E1, elution; M, marker. The eluate was used for assays later without further purification as the non-biotinylated impurities were to be eliminated upon protein immobilization on magnetic streptavidin-functionalized crosslinked agarose. **B)** Calibration standard of the mCherry construct for the calculation of the specific fluorescence of mCherry. Calibration standard acquired by measuring the fluorescence of mCherry for different mCherry concentrations. The specific fluorescence thus determined is 48,529 RFU $\mu M^{-1}$. **C)** Release of immobilized mCherry by TEV. Magnetic crosslinked agarose (MagSi-STA 600 BI, MD21001, Steinbrenner Laborsysteme GmbH) and magnetic macroporous cellulose (High Capacity Magne® Streptavidin Beads, V7820, Promega) were functionalized with biotinylated mCherry as described in the Experimental Section. Fluorescence of mCherry (excitation: 575 nm, emission: 620 nm) determined in the supernatant after incubation of the mCherry construct with streptavidin-functionalized magnetic agarose (FT), after each wash step (W1 – W5), and after elution (E) by TEV (incubation with 7 µM TEV for 2 h at 37 °C).

**Description of the Mathematical model**

**1. Mathematical model of the CRISPR/Cas-based ssRNA detection system**

We developed a mathematical model to characterize, optimize and predict the behavior of a biomaterials-based ssRNA detection system. The model consists of ordinary differential equations (ODEs) describing the dynamic behavior of the relevant system components. Enzymatic reactions were modeled via mass action kinetics and diffusion was approximated by the use of linear chains.

The model scheme of the ssRNA detection system is shown in **Figure S4**. The Cas13a/crRNA complex, named Cas13a-inactive in the scheme, gets activated upon binding of its defined trigger RNA (v1) and deactivates over time (v2). TEV, fused to a Halo-tag, is attached to magnetic beads by streptavidin and a reporter RNA. This reporter RNA gets cleaved by active Cas13a, releasing TEV (v6). Due to the construction of the system, where many beads loaded with TEV constructs are all localized over the same bar magnet, the reporter RNA is surrounded by very dense material and is not directly accessible for Cas13a. To approximate the diffusion of Cas13a through this dense material to the cutting site, we used a linear chain[1] with the two intermediate states $Cas13a_{d1}$ and $Cas13a_{d2}$ and the transitions v3-v5. Once TEV is released, it can either diffuse to the mCherry subsystem, which is also based on coated magnetic beads located over a second magnet, or deactivate (v7). mCherry is genetically fused to an AviTag, which attaches to the magnetic beads, via a linker containing a cutting site for the TEV protease. Similar to the TEV subsystem, mCherry construcst are also packed very densely over the magnet and TEV has to diffuse to its cutting site (v8-v10) to release mCherry (v11) and thereby induce a detectable fluorescence signal. As the two subsystems are not completely separated but constructs might interact from time to time, the bound TEV protease also leads to a release of mCherry to a small extend (v12). In addition an unspecific leaky release of mCherry was observed (v13), which might be caused by the overload of the magnetic beads with mCherry constructs. As mCherry is released, the access to the TEV cutting site gets easier leading to a positive feedback of mCherry on its own release. The model consists of 13 reactions with corresponding fluxes v1-v13 (**Table S3**). Given these fluxes, the resulting set of ODEs can be formulated (**Table S4**) to describe the dynamic behavior of the system.

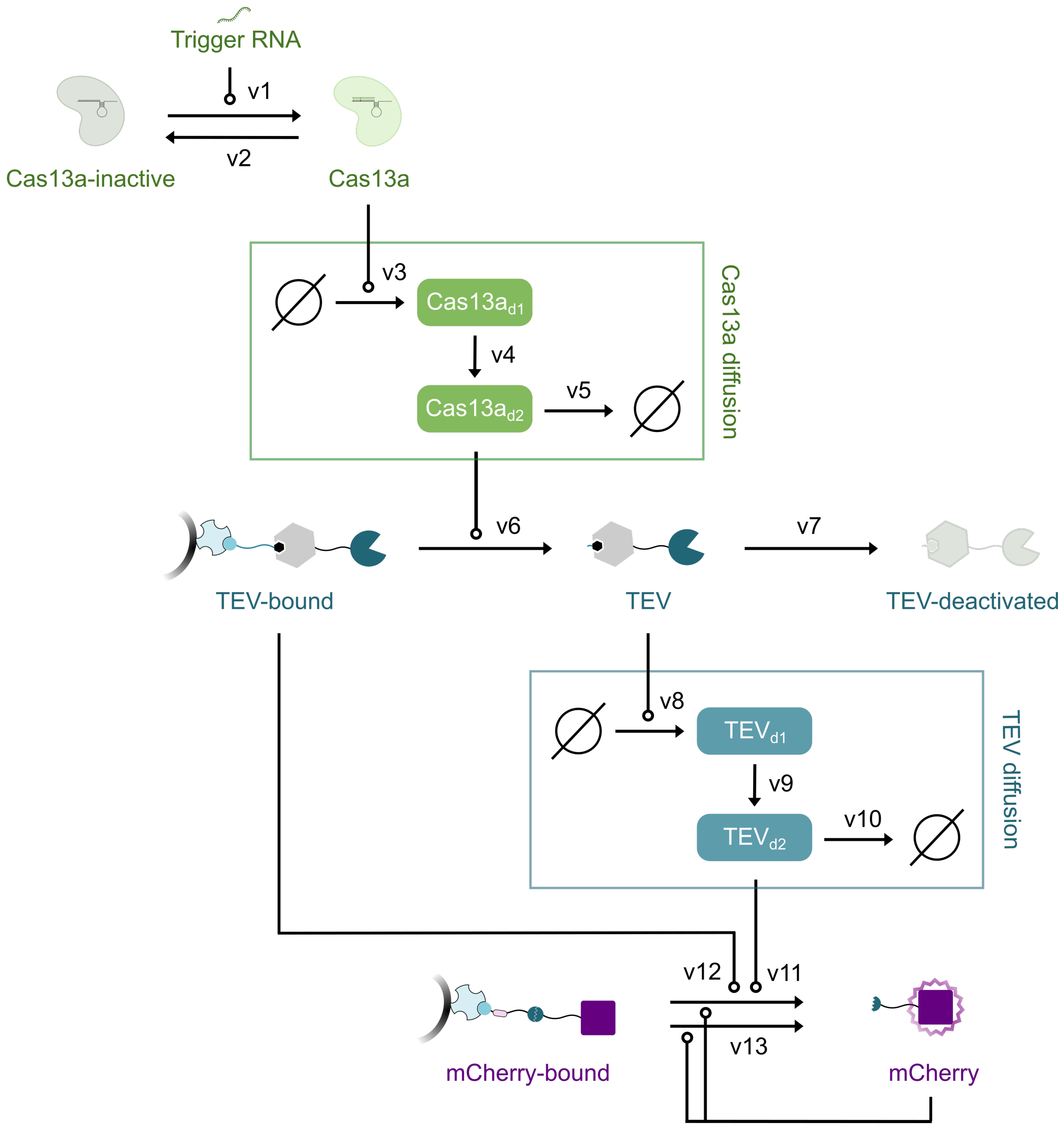


**Figure S4. Model scheme of the ssRNA detection system.** Cas13a is activated by the trigger RNA (v1), can deactivate (v2) and diffuse into the TEV submodule (v3-v5) respectively. TEV-bound is released by the diffused Cas13a (v6), can the get deactivated (v7) or translocate into the mCherry subsystem (v8-v10). mCherry-bound is released by the translocated TEV (v11), by TEV-bound though accidental interaction (v12) or by a leaky release (v13). This mCherry release is enhanced by a positive feedback loop.

### 2. Likelihood-based parameter estimation and identifiability analysis

The parameters of the mathematical model were initially unknown and thus, inferred from experimental data through maximum likelihood estimation. We established a connection between internal model states and experimental data via an observation function. A constant Gaussian error was assumed as measurement noise with its variance estimated concurrently with the dynamic model parameters. To assess parameter uncertainties and consequently the system's identifiability, we computed parameter confidence intervals employing the profile likelihood method.[2] Further, these parameter profiles were utilized to reduce the model, as outlined previously, to enhance the system's identifiability.[3]

### 3. Implementation of single experiments

Three different experiments, characterizing the time- and dose-resolved release of TEV and mCherry, were used to calibrate the mathematical model, extended by a fourth experiment for model validation:

Experiment 1: Characterization of the TEV subsystem

In this experiment, the TEV subsystem was implemented to quantify the release of TEV in dependence of different ssRNA concentrations (**Figure 4B**). Thus, this subsystem captures the activation of Cas13a, its diffusion to the reporter RNA and the release of TEV. The release was monitored over 120 minutes after stimulation with four different concentrations of trigger RNA and without as negative control. The initial concentration of inactive Cas13a was fixed to the applied concentration of 0.05 μM, those for released and deactivated TEV, active Cas13a and the two Cas13a diffusion states were fixed to 0 and the initial of TEV-bound was estimated as model parameter.

Experiment 2: Characterization of the mCherry subsystem

In this experiment, the mCherry subsystem was implemented to quantify the release of mCherry in dependence of different TEV (unbound) input concentrations (**Figure 4C**). Thus, this subsystem captures the diffusion of TEV to the TEV cleavage site in the mCherry construct and the release of mCherry. The release was monitored over 210 minutes after stimulation with three different concentrations of TEV and without as negative control. The initial concentrations of free mCherry, deactivated TEV and the two TEV diffusion states were fixed to 0, while the initial of mCherry-bound was estimated as model parameter.

Experiment 3: Characterization of the whole biohybrid circuit

In this experiment, both subsystems were combined to quantify the release of mCherry in dependence of different ssRNA concentrations (**Figure 4D**). Thus, this subsystem captures all reactions included in the model scheme (**Figure S4**). The release of mCherry was monitored over 210 minutes after stimulation with three different concentrations of TEV and without as negative control. The initial concentration of inactive Cas13a was fixed to the applied concentration of 0.05 µM, while the initials of TEV-bound and mCherry-bound were estimated as model parameters. All other model states were assumed to be not present at the beginning of the experiment at therefore fixed to 0.

Experiment 4: Validation measurements for the whole biohybrid circuit

The setup of this experiment was identical to Experiment 3. However, only two ssRNA concentrations, namely 0 and 0.1 µM, were used for the stimulation of the system and measurements were taken only after 240 minutes. The resulting data was not used for the calibration of the mathematical model but used for validation as described in section 5 “Model predictions and validation”.

**4. Fitting process and results**

The model was first calibrated on the two subsystems before combining them to the complete ssRNA detection system. When combining the two modules, the additional release of mCherry by bound TEV (v12) became necessary, as this interaction was impossible in the separated submodules. All model parameters were simultaneously estimated from the experimental data. In totality, the model encompasses 23 parameters, whereof ten describe dynamic parameters, eleven represent initial concentrations and two are error parameters. To ensure positivity and numerical stability, parameters were subjected to log-transformation.

The compilation, numerical integration, fitting, and optimization of the model were executed using dMod, a freely available software package implemented in R.[4] Deterministic multi-start optimization was conducted employing the trust region optimizer.[5] Among the total of 40 fits performed, 34 converged to the identical lowest minimum, indicating the global optimum. The resultant model curves and associated data are depicted in **Figure 4B-D**. The shaded bands in the figure represent the estimated standard deviation of the Gaussian error model. Estimated parameter values corresponding to the global optimum along with profile likelihood-derived confidence intervals are depicted in **Figure S5** and listed in **Table S5** on the linear scale.

### 5. Model predictions and validation

In order to assess and optimize the amplification of the ssRNA detection system, the model was used to generate predictions of the systems amplification behavior for RNA input concentrations between 0.001 and 0.8 μM over 300 minutes (**Figure 4E**). The amplification, color-coded in the figure, was defined as follows:

$$\text{Amplification}=\frac{\left(\left[\text{mCherry}_{\text{afterRNAstimulation}}\right]-\left[\text{mCherry}_{\text{negativeControl}}\right]\right)}{\max([\text{TEV}])}$$

To validate the model, the mCherry output of the ssRNA detection system was also experimentally quantified for the RNA input 0 and 0.1 μM after 240 minutes. Measurements of this additional experiment of the complete system were in agreement with the model predictions and could thereby validate the model (**Figure 4E**).

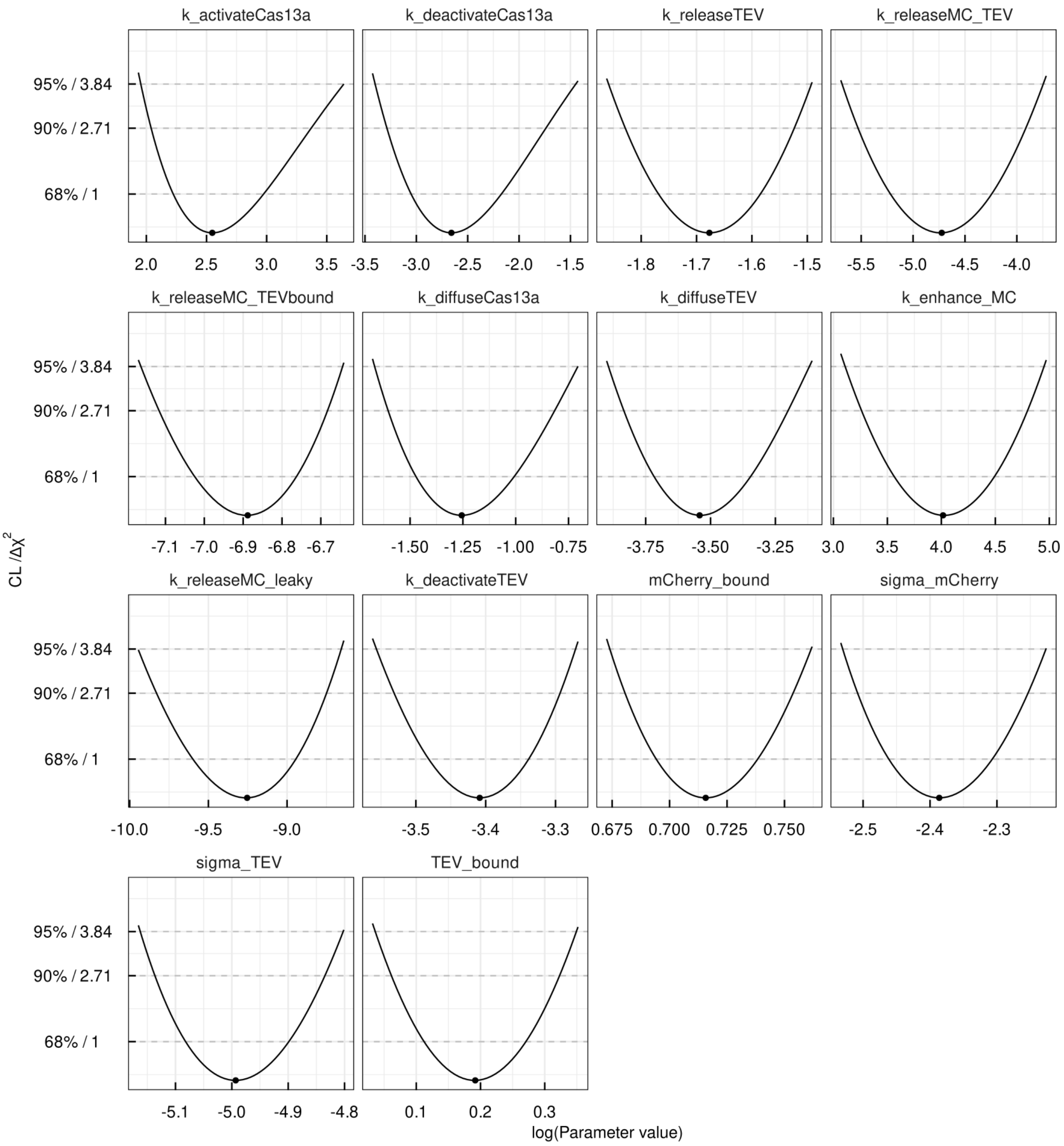


**Figure S5.** Profile likelihood of estimated parameter values. It is represented by continuous lines with a pinpoint marking the optimal parameter value. Confidence levels are illustrated by dotted lines. Parameter values are displayed on a logarithmic scale.

**Table S3.** Reaction fluxes of the mathematical model as indicated in Figure S4.

| Flux | Educt | Product | Rate | Description |
|---|---|---|---|---|
| v1 | $Cas13a_{inactive}$ | Cas13a | $Cas13a_{inactive}$ · k_activate_Cas13a · $RNA_{Input}$ | Cas13a activation |
| v2 | Cas13a | $Cas13a_{inactive}$ | Cas13a · k_deactivate_Cas13a | Cas13a deactivation |
| v3 | | $Cas13a_{d1}$ | Cas13a · k_diffuse_Cas13a | Cas13 diffusion 1 |
| v4 | $Cas13a_{d1}$ | $Cas13a_{d2}$ | $Cas13a_{d1}$ · k_diffuse_Cas13a | Cas13 diffusion 2 |
| v5 | $Cas13a_{d2}$ | | $Cas13a_{d2}$ · k_diffuse_Cas13a | Cas13 diffusion 3 |
| v6 | $TEV_{bound}$ | TEV | $TEV_{bound}$ · k_release_TEV · $Cas13a_{d2}$ | TEV release by Cas13a |
| v7 | TEV | $TEV_{deactivated}$ | TEV · k_deactivate_TEV | TEV deactivation |
| v8 | | $TEV_{d1}$ | $TEV^2$ · k_diffuse_TEV | TEV diffusion 1 |
| v9 | $TEV_{d1}$ | $TEV_{d2}$ | $TEV_{d1}$ · k_diffuse_TEV | TEV diffusion 2 |
| v10 | $TEV_{d2}$ | | $TEV_{d2}$ · k_diffuse_TEV | TEV diffusion 3 |
| v11 | $mCherry_{bound}$ | mCherry | $mCherry_{bound}$ · k_release_MC_TEV · $TEV_{d2}$ · (1 + mCherry · k_enhance_MC ) | mCherry release by free TEV |
| v12 | $mCherry_{bound}$ | mCherry | $mCherry_{bound}$ · k_release_MC_$TEV_{bound}$ · $TEV_{bound}$ | mCherry release by bound TEV |
| v13 | $mCherry_{bound}$ | mCherry | $mCherry_{bound}$ · k_release_MC_leaky · (1 + mCherry · k_enhance_MC ) | mCherry leaky release |

**Table S4.** Ordinary differential equations of the mathematical model.

$$\frac{d}{dt}Cas13a = -\frac{d}{dt}Cas13a_{inactive} = v1 - v2$$

$$\frac{d}{dt}Cas13a_{d1} = v3 - v4$$

$$\frac{d}{dt}Cas13a_{d2} = v4 - v5$$

$$\frac{d}{dt}TEV_{bound} = - v6$$

$$\frac{d}{dt}TEV = v6 - v7$$

$$\frac{d}{dt}TEV_{deactivated} = v7$$

$$\frac{d}{dt}TEV_{d1} = v8 - v9$$

$$\frac{d}{dt}mCherry = -\frac{d}{dt}mCherry_{bound} = v11 + v12 + v13$$

**Table S5.** Estimated parameter values of the mathematical model. Parameter values are determined using maximum likelihood estimation. The upper and lower bounds indicate the 95% point-wise confidence intervals calculated based on the profile likelihood method.

| Parameter | Optimal value | Lower bound | Upper bound |
|---|---|---|---|
| k_activate_Cas13a | $1,28 \cdot 10^{1}$ | $7,05 \cdot 10^{0}$ | $3,82 \cdot 10^{1}$ |
| k_deactivate_Cas13a | $7,00 \cdot 10^{-2}$ | $3,33 \cdot 10^{-2}$ | $2,35 \cdot 10^{-1}$ |
| k_release_TEV | $1,87 \cdot 10^{-1}$ | $1,56 \cdot 10^{-1}$ | $2,25 \cdot 10^{-1}$ |
| k_release_MC_TEV | $8,88 \cdot 10^{-3}$ | $3,40 \cdot 10^{-3}$ | $2,36 \cdot 10^{-2}$ |
| k_release_MC_$TEV_{bound}$ | $1,02 \cdot 10^{-3}$ | $7,75 \cdot 10^{-4}$ | $1,30 \cdot 10^{-3}$ |
| k_diffuse_Cas13a | $2,85 \cdot 10^{-1}$ | $1,89 \cdot 10^{-1}$ | $4,94 \cdot 10^{-1}$ |
| k_diffuse_TEV | $2,90 \cdot 10^{-2}$ | $2,04 \cdot 10^{-2}$ | $4,42 \cdot 10^{-2}$ |
| k_enhance_MC | $5,55 \cdot 10^{1}$ | $2,24 \cdot 10^{1}$ | $1,41 \cdot 10^{2}$ |
| k_release_MC_leaky | $9,60 \cdot 10^{-5}$ | $4,80 \cdot 10^{-5}$ | $1,74 \cdot 10^{-4}$ |
| k_deactivate_TEV | $3,31 \cdot 10^{-2}$ | $2,85 \cdot 10^{-2}$ | $3,80 \cdot 10^{-2}$ |
| $TEV_{bound}$ | $1,21 \cdot 10^{0}$ | $1,04 \cdot 10^{0}$ | $1,42 \cdot 10^{0}$ |
| $mCherry_{bound}$ | $2,05 \cdot 10^{0}$ | $1,96 \cdot 10^{0}$ | $2,14 \cdot 10^{0}$ |
| sigma_mCherry | $9,20 \cdot 10^{-2}$ | $7,96 \cdot 10^{-2}$ | $1,08 \cdot 10^{-1}$ |
| sigma_TEV | $6,78 \cdot 10^{-3}$ | $5,73 \cdot 10^{-3}$ | $8,21 \cdot 10^{-3}$ |

### References


[1] A. L. Hauber, R. Engesser, J. Vanlier, J. Timmer, *Bioinformatics* **2020**, *36* (6), 1848.

[2] A. Raue, C. Kreutz, T. Maiwald, J. Bachmann, M. Schilling, U. Klingmüller, J. Timmer, *Bioinformatics* **2009**, *25* (15), 1923.

[3] T. Maiwald, H. Hass, B. Steiert, J. Vanlier, R. Engesser, A. Raue, F. Kipkeew, H. H. Bock, D. Kaschek, C. Kreutz, J. Timmer, *Plos One* **2016**, *11* (9).

[4] D. Kaschek, W. Mader, M. Fehling-Kaschek, M. Rosenblatt, J. Timmer, *J Stat Softw* **2019**, *88* (10), 1.

[5] C. J. Geyer, trust: trust Region Optimization. 0.1-8 ed.; **2004**.
